# Dynamic Topic Alignment and Sentiment between Official Health Communication and General Public Discourse during COVID-19: A Comprehensive Infoveillance Framework

**DOI:** 10.64898/2026.05.23.26353966

**Authors:** Shuhua Yin, Wangjiaxuan Xin, Yaorong Ge, Shi Chen

## Abstract

Social media has become a critical channel for public health communication during the COVID-19 pandemic, yet how official health messaging aligns with broader public discourse remains insufficiently understood. This study develops an end-to-end infoveillance framework to examine the dynamic relationship between Centers for Disease Control and Prevention (CDC) communications and general public discourse on social media. We analyzed 17,524 CDC tweets and 67,895 public discourse tweets. Biterm Topic Model (BTM) was used to extract topics from each corpus, and a novel topic consistency scoring system integrating cosine similarity with daily public topic prominence was developed to quantify temporal alignment between official health communication and public discourse. Two complementary sentiment measures were incorporated: expected sentiment (average emotional tone) and net sentiment (overall emotional intensity). Temporal relationships were examined using autoregressive integrated moving average with exogenous variables (ARIMAX) models. Results show that topic alignment increased over time across CDC topics, while expected sentiment remained consistently negative. Higher alignment was associated with immediate and delayed changes in expected sentiment and stronger emotional intensity in net sentiment based on ARIMAX results. These findings suggest that topic alignment reflects public attention rather than agreement with official communications and is associated with more negative emotional responses. This framework provides a scalable, generalizable approach to investigate and evaluate public engagement with official health communication.

## 1. Introduction

Public health agencies have been making sustained efforts to inform and guide the public during the COVID-19 pandemic. Social media platforms, where news spread rapidly, contain large-scale and extensive public engagement in real time, and have become essential channels for official health communications. Health agencies, particularly the Centers of Disease Control and Prevention (CDC), have been actively leveraging social media to communicate with the public by sharing epidemic updates, announcing policies and recommendations, and providing guidelines for intervention and prevention strategies throughout the pandemic. At the same time, social media also hosts a massive and evolving landscape of general public discourse, where individuals express opinions, emotions, concerns, and perceptions in real time. It is crucial to evaluate and understand how closely official health communications aligned with the opinions, concerns, and perceptions across the broad public discussions, especially during health emergencies such as COVID-19.

Building on this context, the fields of infodemiology and infoveillance have emerged to utilize large-scale online data for public health insights. Infodemiology (“information” and “epidemiology”), introduced by Gunther Eysenbach, is the study of the distribution and determinants of information in populations through the internet and other digital platforms [1]. Infoveillance, extends this concept by focusing on real-time surveillance, using innovative computational methods to analyze unstructured large scale online data and monitor public health dynamics. Compared to traditional epidemiological surveillance systems, such as cohort studies, registries, surveys, and clinical records, infoveillance approaches enable more timely and scalable analysis by utilizing real-time digital data streams. These methods support detecting trends and patterns, monitoring and assessing public opinions, perceptions, and emotions towards key health policies during health emergencies [2]. As a result, infoveillance provides a more time-efficient approach to traditional surveillance systems, offering a dynamic perspective on how public discourse evolves during public health emergencies.

While existing studies have examined online public sentiment and discussions, there is still a lack of systemic investigation on the dynamic alignment between those focal messages by official health agencies and the evolving topics of the general public discourse in a comprehensive framework that integrates multiple methodologies. Li et al [3] developed a natural language processing (NLP) pipeline to extract pharmacotherapy-related topics and analyze public comments and sentiments on social media during the COVID-19 pandemic. Boukobza et al [4] incorporated deep neural networks and lexical sentiment analysis to capture public topics and sentiments simultaneously from the COVID-19-related tweets posted the day after the World Health Organization (WHO) announced the COVID-19 pandemic on Twitter. Although this approach captured immediate public reactions, the analysis was limited to a single day of data, restricting its ability to reflect continuous public engagement and temporal dynamics. Rao et al [5] examined focal issues and emotional language from public health experts and non-experts during COVID-19 but did not model how topic alignment and emotions evolve over time between public health agencies and a broader general public discourse. Yin et al [2] investigated the dynamic associations between the CDC communication topics on social media and the actual epidemic outcomes during COVID-19 pandemic using topic modeling and time series analysis; however, further work is needed to examine how broader general public discourse and public emotions and align with the official health communication over time, as public opinions heavily influence intervention outcomes.

These limitations highlight the importance of examining temporal dynamics in both public discourse and sentiment. Public opinions and emotions shift in response to various factors, such as events and policy changes, it is critical to understand how public opinions evolve over time and how they align with or diverge from health agencies’ focal communication topics. Therefore, it is important to evaluate the effectiveness of public health messaging during health emergencies. Addressing these limitations helps bridge the gap in understanding how public health communications are received and reflected in the ongoing public discourse, e.g., highly viral or engaged, misinterpreted, or deprioritized. This study aims to investigate the thematic and temporal alignment between CDC communications and general public discourse.

Unlike traditional approaches that focus on direct public responses to institutional communications, this study examines and captures the temporal alignment between CDC communications and a broader, independent general public discourse, providing a more comprehensive view of public engagement and reactions. In this study, we seek to answer the following major research questions:

RQ 1. How do public opinions, expressed as various content topics and sentiments, correlate with focal content topics of CDC health communications during COVID-19?

RQ2: How to effectively quantify the topic alignment or discrepancies between the CDC communications and general public discourse?

RQ3: How does the consistency between general public discourse and official health communications evolve over time?

## 2. Materials and Methods

### Data retrieval and preprocessing

This study is based on two comprehensive Twitter/X datasets on COVID-19 during the pandemic: CDC communication and general public discussions. A total of 17,524 English CDC tweets dataset from December 17, 2019, to January 15, 2022, was retrieved using the Twitter/X academic API (application programming interface) and a set of relevant keyword-based search query. These tweets were posted by 7 official CDC-affiliated Twitter accounts. A total of 67,895 general public discourse tweets, from February 28, 2020, to June 1, 2023, was collected from Twitter/X using the BrandWatch platform [6] and a set of keyword-based search query encompassing medical, colloquial, and politicized references. This dataset provides a foundation for comprehensive downstream analyses of public sentiment, topic content, and engagement patterns over the course of the pandemic.

A series of natural language processing steps were performed to preprocess both datasets in Python. Details of search queries, data collection, and computational linguistic processes are provided in Supplementary Materials.

### Topic Modeling (Biterm Topic Model)

Biterm topic model (BTM) was applied to identify and extract key topics from both CDC communications and general public discourse. BTM is well-suited for short-text data such as Twitter/X posts and other social media textual data [7], as it models word co-occurrence patterns across the entire corpus (i.e., collection of all posts) rather than within individual documents (i.e., a single post). This approach addresses the data sparsity issue of traditional topic models, e.g., latent Dirichlet allocation (LDA), when applied to short texts on social media, enabling robust extraction of semantically coherent topics from large-scale online discourse datasets (e.g., a corpus of many posts) [8]. Additional technical details are provided in Supplementary Materials for BTM.

In this study, BTM was applied separately to the CDC communication corpus and the general public discourse corpus on COVID-19 using the *bitermplus* package in Python [9]. Modeling the two corpora separately allows each corpus to retain its own topic structure, reflecting differences in communication style, focus, tone, and language use between official health communication and public discussions during the pandemic. As a result, two independent and distinct sets of topics were derived: CDC topics and general public discourse topics representing themes in public discussion and engagement. A unique shared vocabulary was constructed for comparability between the two corpora. Each topic was represented in a probability distribution over this shared vocabulary, allowing direct comparison of topic-word distributions across the two corpora.

To determine the optimal number of topics and the corresponding topic themes for each corpus, we used a hybrid quantitative-qualitative approach. BTM models were evaluated across a range of topic numbers (5 to 12) using Rényi entropy [10] (see Supplementary Materials for details). Lower entropy values indicate more cohesive and well-separated topic distributions [11]; therefore, the number of topics was selected based on a local minimum followed by stabilization in the entropy curve. For each topic, 20 randomly sampled tweets, along with the top associated keywords and visualizations outputs of the topic clusters generated by BTM, were examined. Domain experts iteratively assessed these topic clusters for interpretability and thematic coherence. The optimal number of topics and topic themes were adjusted and refined through repeated expert evaluation until consensus was reached.

### Quantifying Topic Alignment across corpus: the Topic Consistency Scoring System

A key objective of this novel infoveillance framework is to quantify the dynamic alignment or divergence between topics from the CDC health communication and general public discourse during the COVID-19 pandemic. To achieve this, we developed a novel topic consistency scoring system that integrates both semantic similarity and the daily prominence of public discourse topics, capturing both topic alignment and shifts in public focus over time. This system is based on a weighted similarity formulation that compares topic-word probability distributions derived from the two independent corpora: CDC communications and general public discourse topics.

To measure the semantic alignment, we computed pairwise similarity between CDC topics and general public discourse topics based on their topic-word probability distributions derived from the BTM [9]. Each topic is represented as a probability distribution over a shared vocabulary. Cosine similarity was used to quantify the similarity between these topic vectors [12], capturing how closely the semantic content of general public discourse aligns with the CDC communication at a given time. These pairwise similarities between topic-word distributions of the two corpora were computed using the cosine similarity function in *scikit-learn* package [13] in Python (See Supplementary Materials for detailed information on cosine similarity). However, semantic similarity alone does not account for how prominently each topic appears in public discourse on a given day. Although the cosine similarity matrix provides pairwise similarities between individual CDC and public discourse topics, it is not used for one-to-one topic matching. To address this, daily topic prominence was incorporated as weights across all general public topics. In this way, the cosine similarity matrix defines possible associations between topics, while the weighted system determines how these relationships contribute to overall alignment on each day.

More specifically, these pairwise cosine similarities are aggregated using daily topic prominence weights to construct a weighted alignment score that reflects the overall public discourse relative to each CDC topic. These weights were calculated as the proportion of each general public topic on a given day, relative to the total number of general public discourse for that day. The final weighted sum of similarities, or topic-level consistency, across all public discourse topics, for CDC topic *j* on day *d* is defined as: 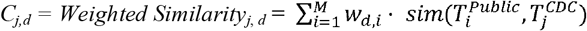, where *d*: index of the day in the daily general public discourse time series

*i* = 1, …, *M*: general public topic index

*j* = 1, …, *N*: CDC topic index

w_*d,i*_: weight of public topic *i* on day *d*, computed as:

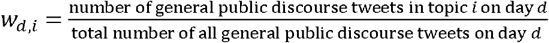

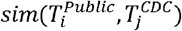 cosine similarity between the topic-word distribution probability matrices of general public topic *i* and CDC topic *j*.

Because the CDC topic-word distributions are treated as fixed reference topics, the daily topic consistency score can be computed across the complete general public discourse timeline. This results in a time series of scores that reflect how general public discourse aligns with these CDC communication topics over time. After the CDC tweet collection period ends, the score is interpreted as the level of alignment between ongoing public discourse and the fixed CDC topics, rather than as alignment with newly posted CDC communications.

Specifically, cosine similarity resulted in an *m* × *n* similarity matrix *S* ∈ ℝ^*m*×*n*^, where each value represents the semantic similarity between a general public topic and a CDC topic. For each day *d*, the weight *w*_*d,i*_ for public topic *i* was defined as proportion of tweets of each topic on that day. This results in a weight matrix of *d* × *m, W* ∈ ℝ^*d*×*m*^, representing the temporal prominence of each topic. The final daily topic consistency scores measure how closely general public discourse aligns with each CDC communication topic over time. Higher values indicate stronger alignment between public discourse and CDC communication topics. This scoring system forms the foundation for the subsequent analyses examining the relationship between topic alignment and public sentiment dynamics.

### Integration of Public Sentiments into Topic Consistency Scoring System

While topic consistency captures the degree of alignment between CDC communication and general public discourse topics, it does not reflect the emotional context in which the alignment occurs. To address this issue, we further incorporated sentiment analysis into the infoveillance framework to examine how public sentiment evolve alongside topic alignment over time during the COVID-19 pandemic. Specifically, we considered and evaluated two complementary measures of public sentiment: (1) net public sentiment, which captures overall emotional intensity, and (2) expected public sentiment, which captures the average emotional tone. These two measures provide distinct but related insights to capture public sentiment. Net sentiment captures the aggregate emotional polarity (difference between positive and negative sentiments), and the overall emotional intensity, or magnitude from the general public towards a topic, thus it is sensitive to the topic prominence. In contrast, expected sentiment is normalized by the total number of posts per day and therefore not influenced by fluctuations in topic prominence. It reflects the average emotional tone towards a randomly selected post within a topic regardless the overall emotion intensity of all the posts on that day. As a result, there may be two time periods with similar average tone but substantially different emotional intensity from the public towards a topic.

To quantify these sentiment dynamics, sentiment scores of the general public discourse were assigned at post level for each tweet. Let s_*d,i,k*_ denote the sentiment score of tweet *k* in public topic *i* on day *d*, where s_*d,i,k*_ = 1 for positive, 0 for neutral, and -1 for negative sentiment, following standard sentiment encoding approaches in social media text analysis [14]. Then the net sentiment of a topic *i* on day *d* is define as:

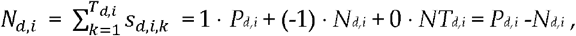

where *P*_*d,i*_, *N*_*d,i*_, and *NT*_*d,i*_ represent the number of positive, negative, and neutral tweets, respectively, and *T*_*d,i*_ = *P*_*d,i*_ + *N*_*d,i*_ + *NT*_*d,i*_ is the total number of tweets for topic *i* on day *d*.

The daily net sentiment across all general public topics is defined as:

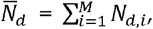

where *M* is the total number of general public topics. This measure captures the overall magnitude and polarity of public emotional expression and reflects how strongly sentiment is concentrated on a given day.

In contrast, the expected daily sentiment of a topic *i* on day *d* is defined as the normalized average:

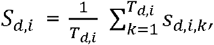

Since *S*_*d,i*_ is a normalized average, it is further weighted by topic prominence to represent the relative percentage of each topic in overall public discourse on a given day, therefore the final daily expected sentiment across all topics is is computed as a weighted sum:

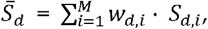

where 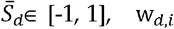 denotes weight and represents the topic prominence of public topic *i* on day *d*, and is computed as:

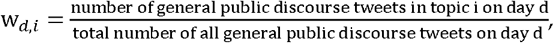

Weights are not applied to net sentiment, which is already a summation of all sentiments, and thus it is a measure of magnitude and sensitive to public discourse volume on a given day.

Together, these two sentiment measures provide complementary and comprehensive insights into public reactions and emotions across different public discourse topics: expected sentiment captures the average emotional tone, while net sentiment captures the strength and concentration of emotional reactions. Finally, topic alignment measures were categorized using tertile-based discretization [15] (33^rd^ and 67^th^ percentiles) into low, moderate, and high levels to enhance interpretability.

### Topic-Sentiment Dynamic System: Integration of Topic Consistency and Multivariate Time Series Analysis

To examine how topic consistency is associated with and/or influenced by general public sentiment over time, we further developed a topic-sentiment dynamic system by integrating our novel topic consistency scoring metric into a multivariate time series model. Specifically, an autoregressive integrated moving average model with external variables (ARIMAX) was applied to model the temporal dynamics between expected public sentiment and topic consistency as the exogeneous input (X).

Autoregressive integrated moving average (ARIMA) consists of autoregressive (AR) [16], moving average (MA) [17] terms, and an order *d* differencing term. The auto-regressive term,

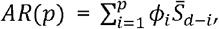

captures the past sentiment values relate to current sentiment (i.e., autocorrelation). The moving average term,

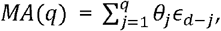

represents the unexplained variation in the sentiment time series. These three components form the *ARIMA*(*p,d,q*) model [18]. See Supplementary Materials for details on autoregressive (AR), moving average (MA) components, and autoregressive integrated moving average (ARIMA) model.

The topic consistency scoring system acts as the independent external variable in the ARIMAX model, forming the final topic-sentiment dynamic system defined as:

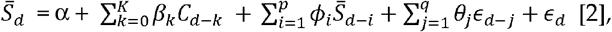

where 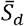, the dependent variable, denotes the expected public sentiment on day *d. C*_*d*−*k*_, the exogeneous independent variable in the ARIMAX model, represents the topic consistency at lag *k*, i.e., the CDC-public topic consistency alignment observed for *k* days prior to day *d*; *β*_*k*_ captures the association between lagged topic consistency and current average public sentiment. Thus,

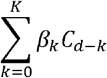

represents the cumulative effects of topic consistency over *K* lagged days. *α* denotes the intercept, and *ϵ*_*d*_ denotes the error term at day *d*.

To complement expected public sentiment, we further modeled net public sentiment in another ARIMAX model. Unlike expected sentiment, which captures average emotional tone, net sentiment reflects the aggregate magnitude and imbalance between positive and negative emotions (i.e., the imbalance between positive and negative sentiment), making it more sensitive to concentrated emotional surges. The final topic-sentiment dynamic system for net public sentiment is defined as:

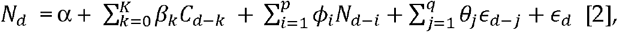

where *N*_*d*_ denotes the net public sentiment on day *d*.

For the ARIMAX models, we chose lag terms up to 7 days to capture immediate and short-term temporal dynamics, potential weekly patterns in public sentiment dynamics, as well as for model interpretability. The optimal values of *ARIMA*(*p,d,q*) model parameters were selected using auto_arima from *pmdarima* package [19] in Python. ARIMAX models were implemented using statsmodels [20] in Python. This dual-model system allows us to distinguish between changes in average emotional tone and shifts in overall emotional intensity of general public sentiment.

## 3. Results

### 3.1. Topic Modeling Results

Datasets of tweets from general public discourse on COVID-19 during the pandemic were collected from BrandWatch platform using keyword-based search queries. A total of 67,895 tweets throughout the pandemic, from February 28, 2020, to June 1, 2023. Biterm Topic Model (BTM) was applied to identify key topics within this corpus. The optimal number of topics was determined by a series of quantitative and qualitative steps. First, Rényi entropy values were computed across a range of topic numbers to identify a local minimum, as shown in Figure 1. Second, topic clusters were examined using visualizations generated by BTM using *tmplot* package [21] in Python, as shown in Figure 2. Finally, domain experts reviewed randomly sampled tweets within each topic to assess interpretability and thematic coherence.

**Figure 1.**
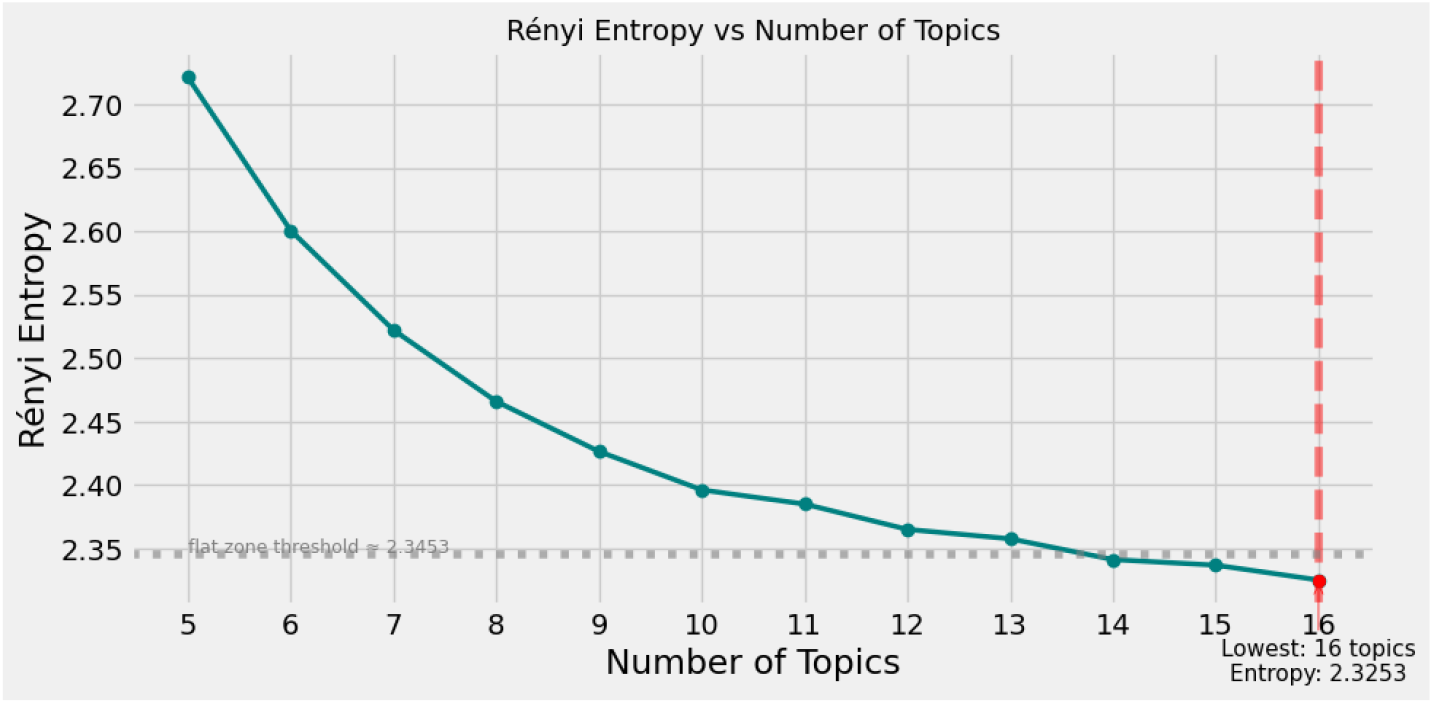
Rényi entropy across different numbers of topics for the Biterm Topic Model (BTM). Lower values indicate more cohesive topics. The curve decreases and begins to stabilize around 10 topics, thus the optimal number of topics for general public discourse.

**Figure 2.**
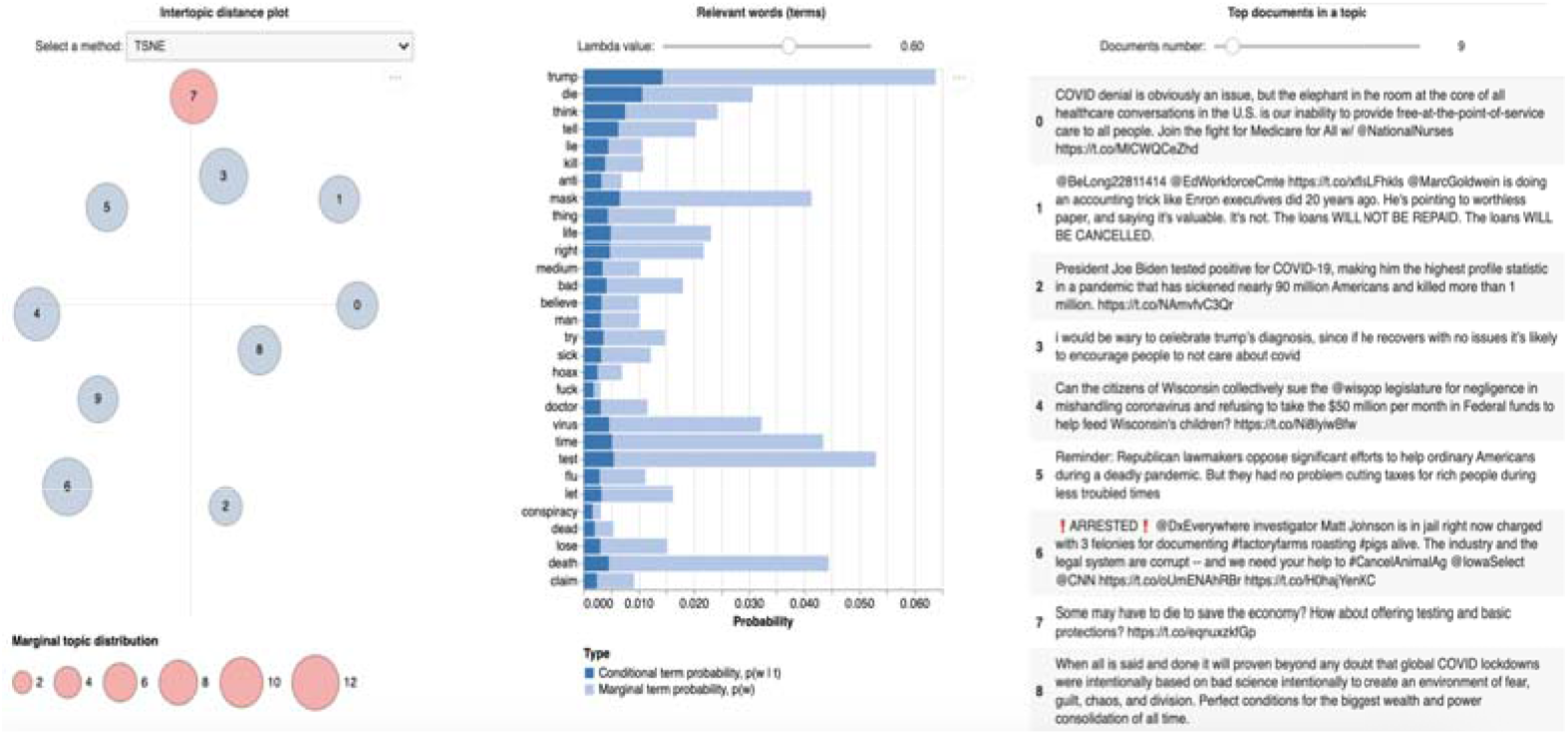
Visualization of Topic 8 (highlighted in pink circle in the left chart) of general public discourse, showing keyword probabilities (middle chart), and top 9 representative sample documents (right chart). Circles represent the topic clusters, size indicates the prevalence of topic in the corpus Topic indices begin at 0, thus this topic appears as topic 7 in the plot. Additional topic visualizations are provided in Supplementary Materials (Figure S2 – S11).

As shown in Figure 1, Rényi entropy decreases as the number of topics increases and begins to stabilize after 10 topics. Therefore, 10 topics were selected as the optimal number, representing a balance between topic coherence and interpretability. The themes of the identified topics and their top 15 associated keywords are provided in Table 1. The same procedure was applied to the CDC communication corpus. 9 CDC topics were identified. The themes of the topics and their top 15 associated keywords are provided in Table 2.

**Table 1.**
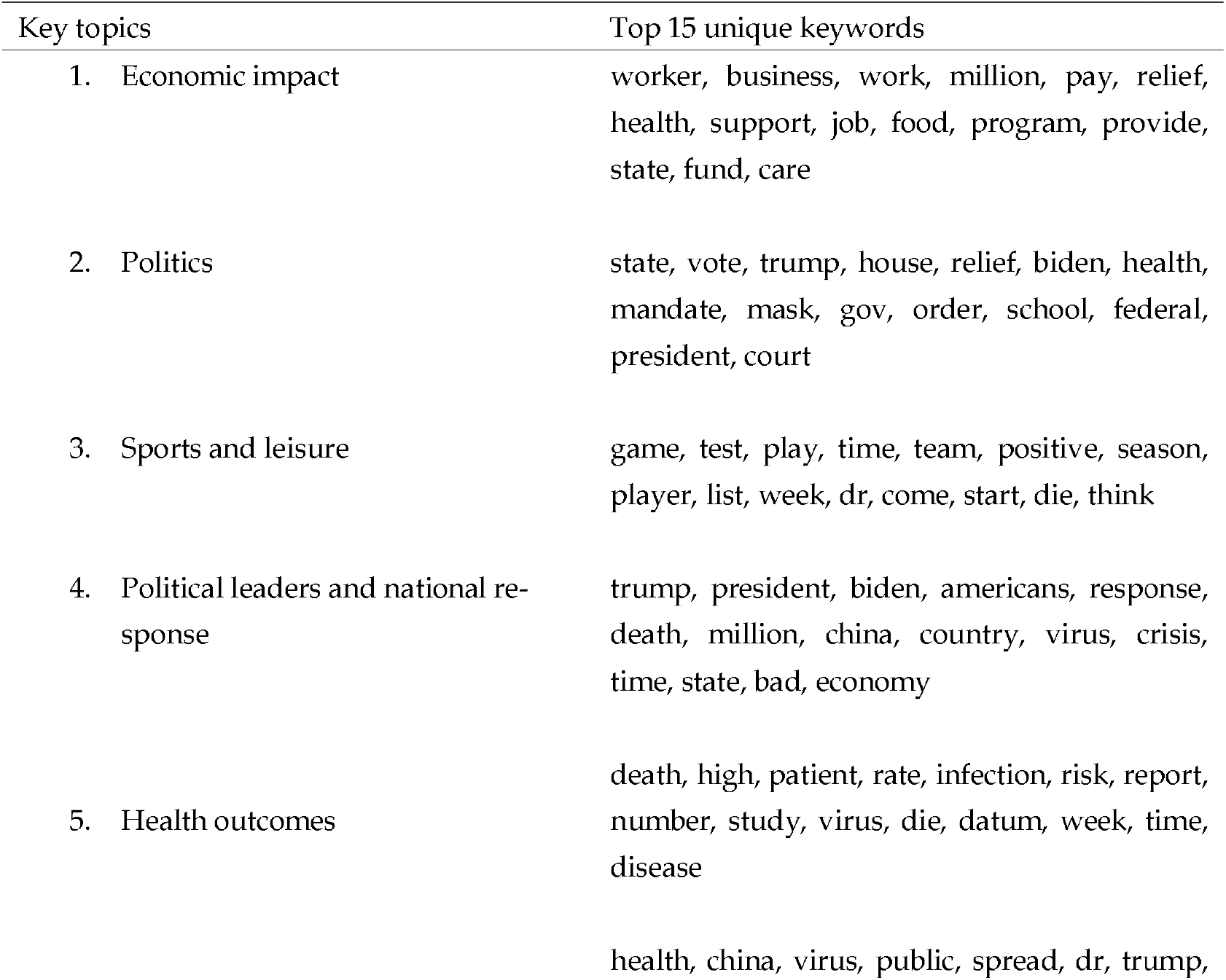
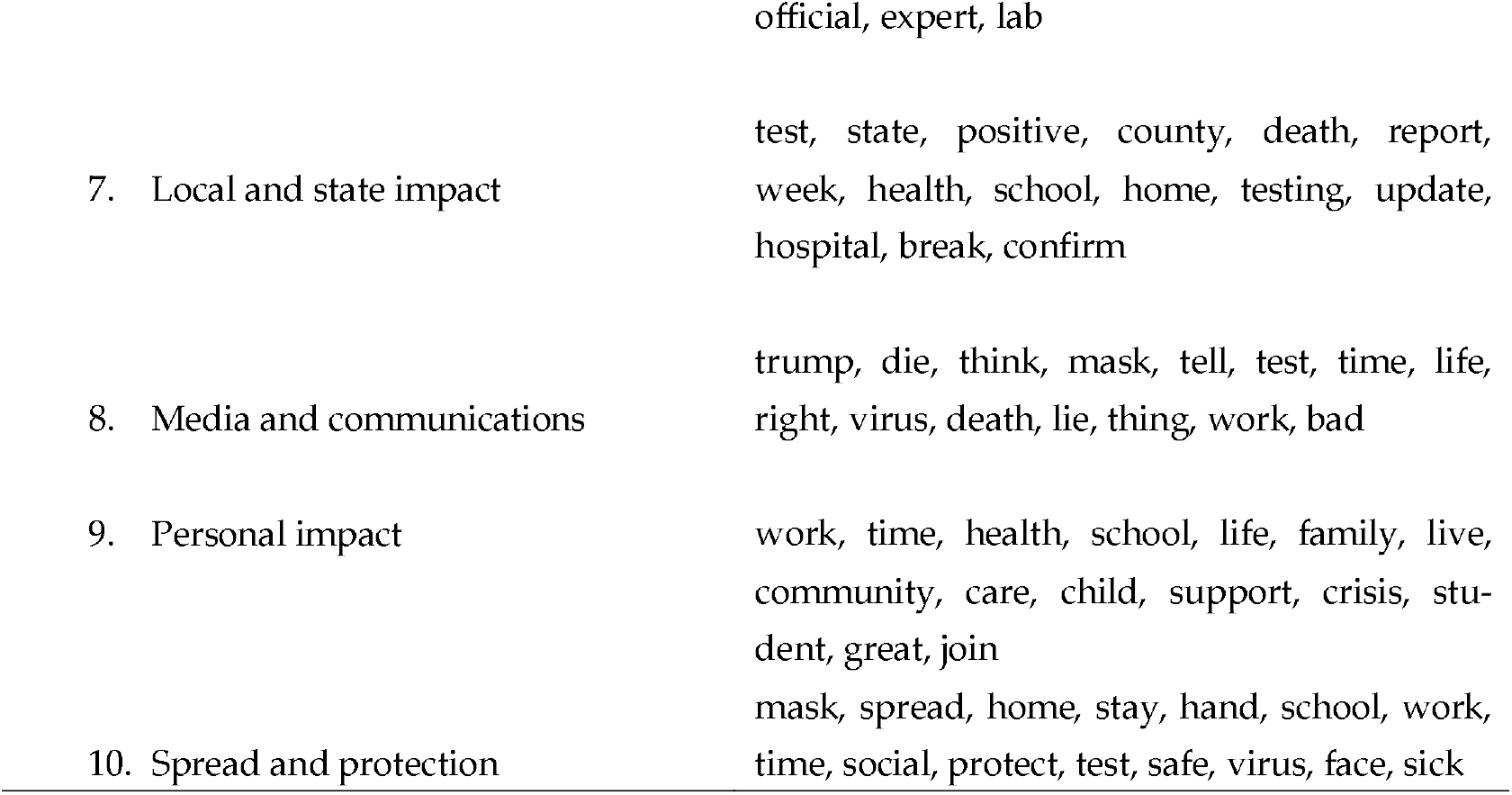
Identified general public discourse topics and their top 15 associated unique keywords.

**Table 2.** Identified Centers for Disease Control and Prevention (CDC) communication topics and their top 15 associated unique keywords.

| Key topics | Top 15 unique keywords |
| --- | --- |
| 1. Epidemic risk and outcome metrics (hospitalization, death) | deport, death, high, increase, rate, risk, flu, week, hospitalization, late, adult, illness, severe, likely, vaccination |
| 2. National and global public health response | hiv, disease, work, public, global, country, prevent, world, partner, program, respond, response, polio, life, support |
| 3. Local and state responses in school and communities | school, test, local, child, community, testing, vaccination, emergency, information, safe, way, protect, follow, spread, prepyourhealth |
| 4. Family protection (prevention and intervention) | protect, spread, flu, vaccinate, virus, wear, family, travel, risk, sick, stay, illness, fully, prevent, important |
| 5. High-risk populations | child, dose, vaccination, safe, receive, year, booster, million, old, age, protect, fully, recommend, shot |
| 6. Healthcare personnel, services, and programs for pandemic response | public, community, work, healthcare, response, dr, program, disease, worker, service, join, spread, partner, support, care |
| 7. Travel and other related health issues during pandemic | travel, emergency, prepyourhealth, family, plan, safe, way, food, stay, like, pet, prepare, hurricane, check, home |
| 8. Non-pharmaceutical interventions | spread, wear, stay, hand, wash, foot, slow, face, avoid, protect, home, cover, apart, cloth, public |
| 9. Clinical aspects and healthcare system | update, patient, pm, et, infection, report, coca, join, topic, healthcare, read, protect, late, hcp, cause |

### 3.2. Topic Consistency Results

A cosine similarity matrix of 10 9 dimension was first computed between the 10 public discourse topics and the 9 CDC communication topics. Each value in this matrix represents the pairwise semantic similarity between a public discourse topic and a CDC topic based on their topic-word probability distributions. Figure 3 shows the heatmap of these similarities, where rows correspond to public discourse topics and columns correspond to CDC topics. Color intensity indicates the degree of cosine similarity, or semantic alignment, between topics.

**Figure 3.**
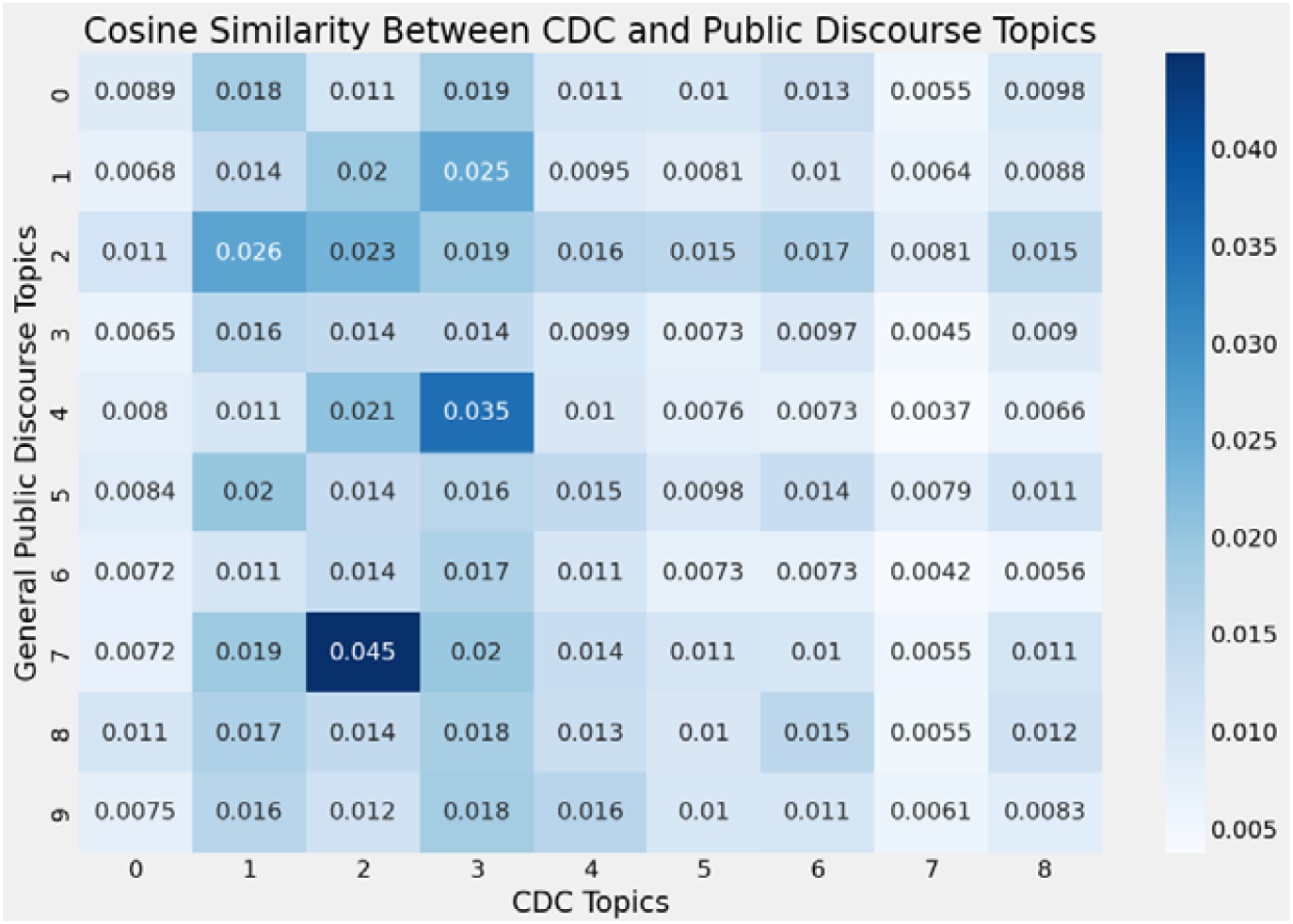
Cosine similarity heatmap between general public discourse topics and the CDC topics. Topic indices start from 0 in the heatmap.

The heatmap shows that certain CDC topics share stronger semantic similarity with specific public discourse topics. For example, CDC Topic 3 (local and state responses in school and communities) shows relatively higher cosine similarity with Public Topic 8 (media and communications), suggesting overlap in semantic themes and language. Similarly, CDC Topic 4 (family protection) demonstrates strong similarity with Public Topic 5 (health outcomes), indicating that discussions around family protection are often expressed in terms of health-related impacts, that the public often viewed family protection in the context of major health outcomes such as deaths and other health impact. In contrast, some CDC topics show consistently lower similarity across public discourse topics. For example, CDC Topic 1 (epidemic risk and outcome metrics such as hospitalization, death) and Topic 8 (non-pharmaceutical interventions) show weaker semantic similarity scores overall. This reflects differences in use of language, where CDC communications exhibit more on technical or specialized and complex health-domain terminology during the pandemic, which were less frequently used in daily general public discussions indicating a noticeable discrepancy between official health communication and public engagement. CDC Topic 8 (non-pharmaceutical interventions) shows the lowest semantic alignment with Public Topic 5 (health outcomes) among all CDC-Public topic pairs, indicating differences in topic prominence, where CDC communications focus more on preventative actions and future health outcomes, while general public discourse largely focuses on current health outcomes such as cases and death rates. These differences in language use and framing are associated with low sematic similarity due to reduced content alignment between these topics.

Cosine similarity provides useful insights into semantic relationships in topic alignment, such as consistency or mismatches in language use and information framing, quantifying content alignment between official health communication and public discourse. However, it only measures semantic (words or languages) alignment and does not account for how prominently each topic appears in general public discourse over time. Therefore, it captures potential content alignment in language and themes but does not reflect the temporal dynamics of public attention over the course of the pandemic.

To address this limitation, daily topic prominence was incorporated to compute weighted similarity scores. These weights were defined as the relative proportion of each general public discourse topic on a given day. The final weighted similarity matrix 1190 9) represents daily topic consistency scores across 1190 days of general public discourse for 9 CDC communication topics. Each value in the matrix represents how general public discourse aligns with a CDC topic on a given day, integrating both semantic similarity and daily public topic prominence.

Figure 4 [22] shows the smoothed daily weighted similarity scores (7-day moving average) for each CDC topic over time. These results demonstrate that topic alignment varies across CDC topics and evolves throughout the pandemic. Some CDC topics exhibit consistently higher alignment with general public discourse, while others remain lower or more stable over time. For example, CDC Topic 3 (local and state responses in schools and communities) and CDC Topic 4 (family protection, prevention, and intervention) show consistently higher weighted similarity scores over time, indicating that these topics are more prominently and strongly reflected in general public discussions. In contrast, other CDC topics’ weighted scores remain relatively low, suggesting their reduced prominence in general public discussions. Figure 5 [22] highlights the highest and lowest alignment trends across CDC topics, illustrating the differences in how strongly individual CDC topics are reflected in general public discourse.

**Figure 4.**
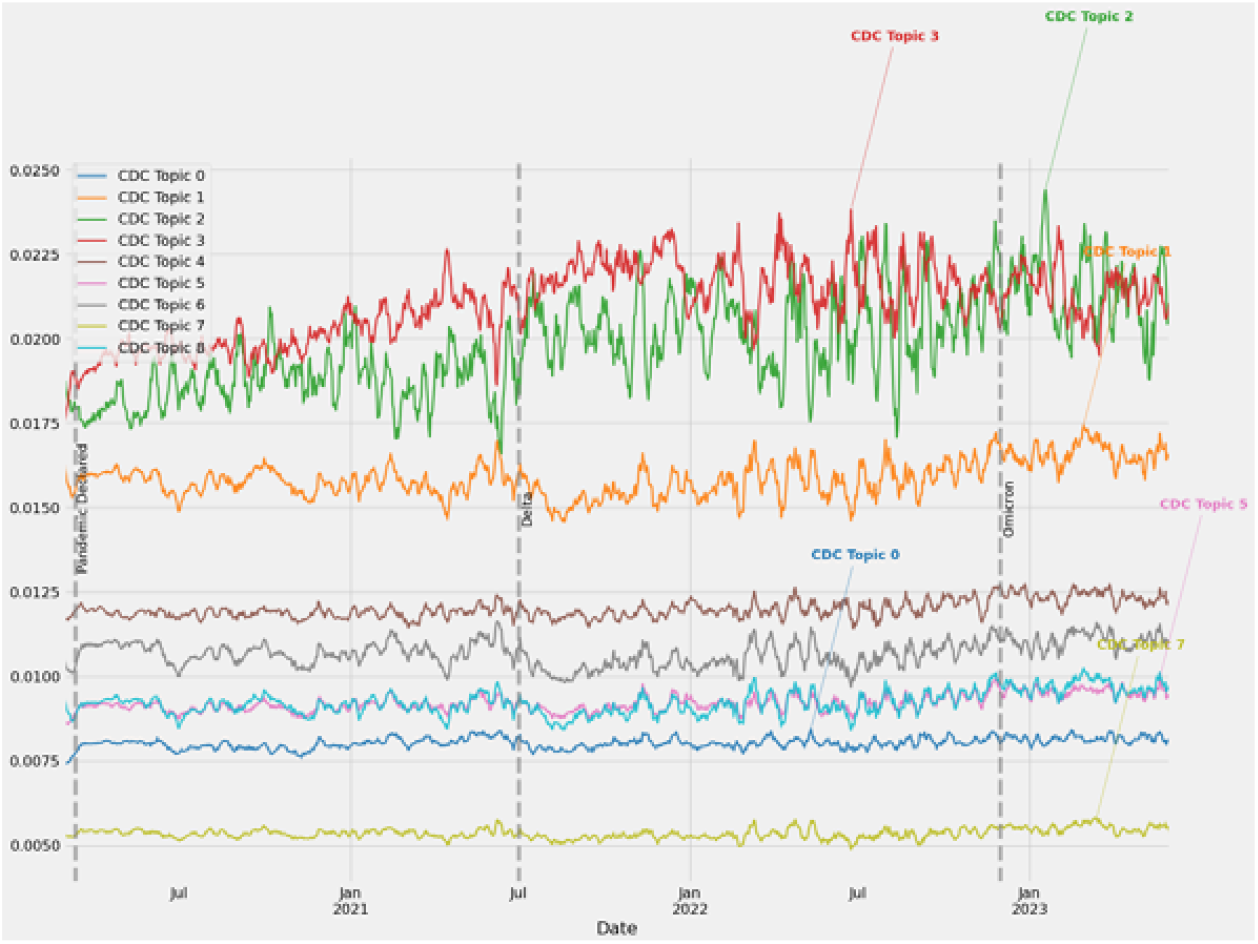
Dynamic topic consistency scores (7-day moving average) between general public discourse topics and the CDC topics.

**Figure 5.**
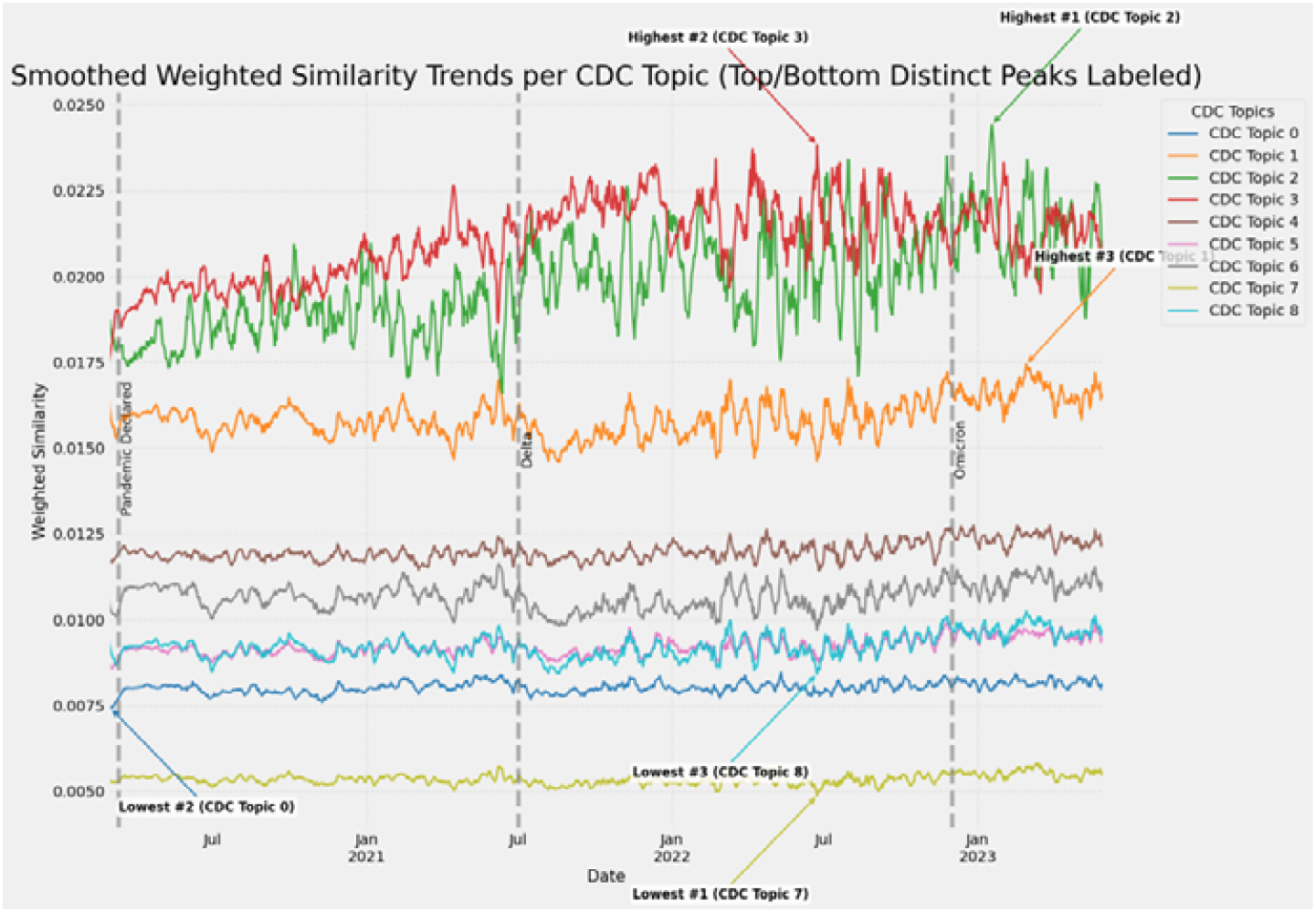
Dynamic weighted similarity scores (7-day moving average) between general public discourse topics and the CDC topics.

Overall, the weighted similarity results capture how general public discourse aligns with CDC communication topics over time by incorporating both semantic similarity and public topic prominence. This dynamic relationship forms the basis for subsequent analyses linking topic consistency with public sentiment measures.

### 3.2. Topic Consistency and Public Sentiment

#### 3.2.1. Topic-Level Alignment Dynamics

We further examined and analyzed how topic alignment evolves alongside expected public sentiment at the topic level. Figure 7 [22] illustrates the topic-level alignment trends together with integrated expected public sentiment across all CDC topics. indicating that public discussions increasingly align with CDC communication topics. However, this increasing alignment does not reflect improvements in public sentiment, which remains consistently negative across topics throughout the study period.

The strength and stability of topic alignment also vary across the CDC topics. For example, CDC Topic 3 (local and state responses in schools and communities) and Topic 4 (family protection and intervention) exhibit consistently strong alignment over time. In contrast, CDC Topic 6 (healthcare personnel and services) and Topic 8 (non-pharmaceutical interventions) show lower alignment over time. In addition, the volatility of this temporal alignment differs across topics. Some display stable and gradual increases in alignment, while others exhibit more abrupt fluctuations across pandemic phases. These patterns suggest that the temporal alignment between CDC communication and public discourse evolves differently across all topics. As shown in Figure 6 [22], alignment between CDC Topic 4 (family protection and intervention) and general public discourse increases steadily over time, but the expected public sentiment remains persistently negative. Moreover, time periods where there has been increasing topic alignment also coincided with decline in sentiment, indicating that stronger alignment could be driven by heightened concern or negative public emotions rather than agreement or positive reception of CDC communications from the public.

**Figure 6.**
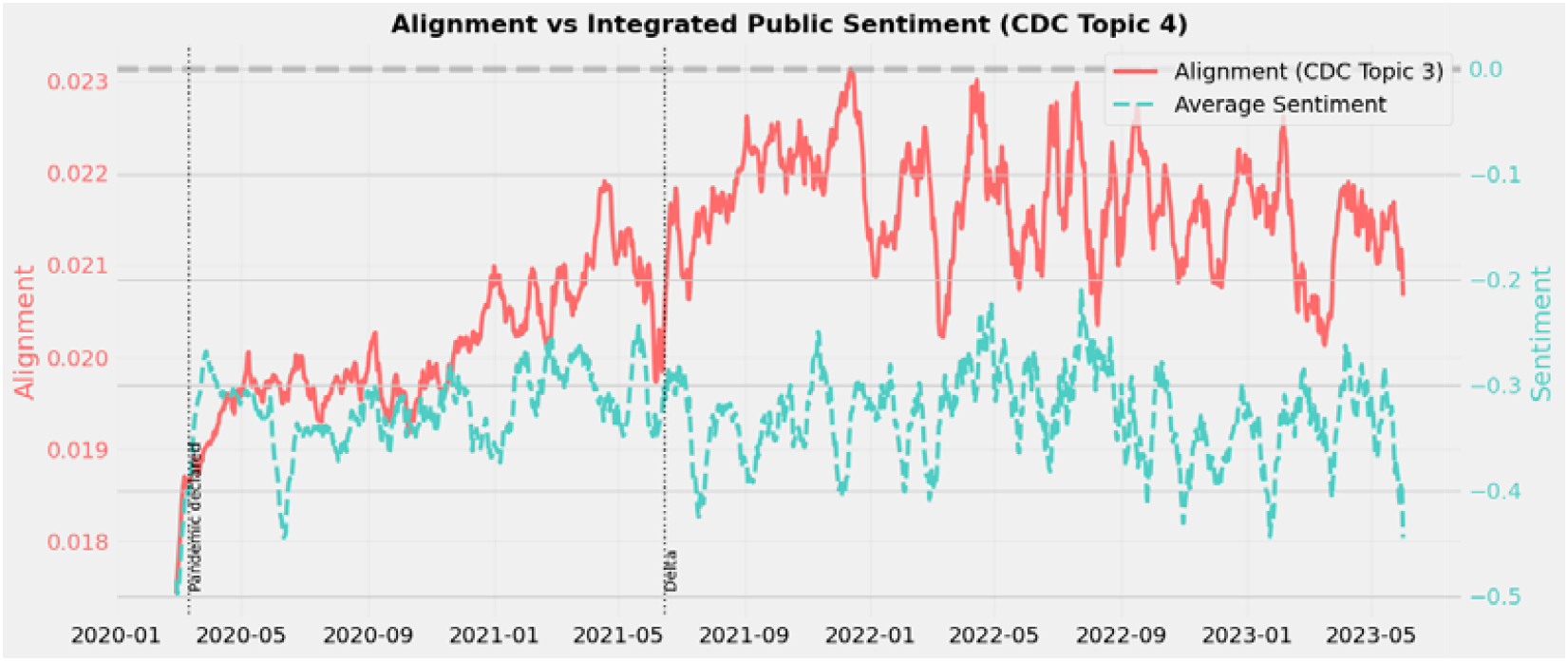
Topic-level alignment and integrated expected public sentiment for CDC Topic 4 over time. Solid line: how consistently public discourse aligns with CDC Topic 4 over time; dashed line: integrated expected public sentiment across topics

Table 3 summarizes these topic-level dynamics. Alignment varies across CDC topics over time. Some topics show consistently higher and more stable alignment, while others show lower alignment or more variable over time. Across all topics, expected public sentiment remains negative even when topic alignment increases, indicating that strong topic alignment reflects public attention and concerns rather than positive reception of official health communications during the pandemic.

**Table 3.** Summary of CDC topic-level alignment patterns.

| CDC Topic number | Key topics | Mean Alignment | Temporal Trend | Volatility | Sentiment |
| --- | --- | --- | --- | --- | --- |
| 1 | Epidemic risk and outcome metrics | Low-Moderate | Gradual increase | Moderate | Negative |
| 2 | National and global public health responses | Moderate | Late increase | Moderate | Negative |
| 3 | Local and state responses in schools and communities | High | Strong increase | High | Negative |
| 4 | Family protection & intervention | High | Sustained increase | Moderate | Negative |
| 5 | High-risk populations | Moderate | Late increase | Moderate | Negative |
| 6 | Healthcare personnel, services & programs | Low | Late increase | Moderate | Negative |
| 7 | Travel & related health issues | Moderate | Fluctuating pattern | High | Negative |
| 8 | Non-pharmaceutical interventions | Low | Slight increase | Low-Moderate | Negative |
| 9 | Clinical aspects & healthcare system | Low-Moderate | Late increase | Moderate | Negative |

#### 3.2.2. Temporal Effects and Dynamics

We distinguish between the two sentiment measures: (1). Expected public sentiment 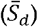, representing the average emotional tone, and (2). Net public sentiment *N*_*d*_, capturing the overall emotional magnitude or intensity.

##### Expected public sentiment 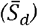

To examine the temporal effects of CDC-public topic consistency on expected public sentiment, we applied a multivariate time series ARIMAX model incorporating lagged topic consistency as the independent exogeneous variable (X), autoregressive (AR), or autocorrelation, of past sentiments, and moving average (MA) effects of random noises or fluctuations in the sentiment time series. The results are displayed in Table 4 as below.

**Table 4.** ARIMAX Results for Expected Public Sentiment with Lagged Topic Consistency.

| Lag (days) | Coefficient ($\beta$) | Standard Error (SE) | z-score | p-value |
| --- | --- | --- | --- | --- |
| 0 | -4.21*** | 0.83 | -5.06 | <0.001 |
| 1 | -0.67 | 0.88 | -0.76 | 0.445 |
| 2 | -2.68** | 0.94 | -2.86 | 0.004 |
| 3 | -1.92* | 0.93 | -2.07 | 0.038 |
| 4 | -0.01 | 0.94 | -0.01 | 0.995 |
| 5 | -0.95 | 0.89 | -1.08 | 0.281 |
| 6 | 0.37 | 0.86 | 0.43 | 0.670 |
| 7 | -5.17*** | 0.86 | -6.03 | <0.001 |

\*\*\*p < 0.001; \*\*p < 0.01; \*p < 0.05
Significant effects observed at lags 0, 2, 3, and 7-day, indicating both immediate and delayed associations between topic consistency and expected public sentiment.

The optimal ARIMAX model order was ARIMAX (4, 0, 4), determined using the auto-ARIMA function in Python, indicating four autoregressive (AR) terms and four moving average (MA) terms with no differencing on the time series data. The ARIMAX model revealed that topic consistency is significantly associated with temporal changes in expected public sentiment. Specifically, increases in topic consistency were strongly associated with immediate, or same day, decreases in the average public sentiment (β = -4.21, p < 0.001). In addition, significant lagged associations were observed at 2- and 3-day lag, indicating that the influence of CDC-public topic consistency is delayed over a few days instead of occurring immediately. Notably, a strong association appeared at a 7-day lag (β = -5.17, p < 0.001), suggesting a potential weekly pattern in expected public sentiment dynamics. This indicates that public sentiments to the topic alignment between CDC communication and general public discourse potentially follow a weekly cyclical pattern. The consistently negative coefficients suggest that higher topic consistency is associated with more negative expected public sentiment, i.e., when the general public discourse closely reflects CDC communication, the overall emotional tone tends to be more negative.

Together, these findings show that the relationship between CDC topic consistency and average public sentiment has immediate, delayed, and recurring patterns of public sentiment dynamics. These temporal dynamics are further illustrated by topic-level patterns in Figure 7, where increases in topic alignment across multiple CDC topics accompany persistently negative expected sentiment.

**Figure 7.**
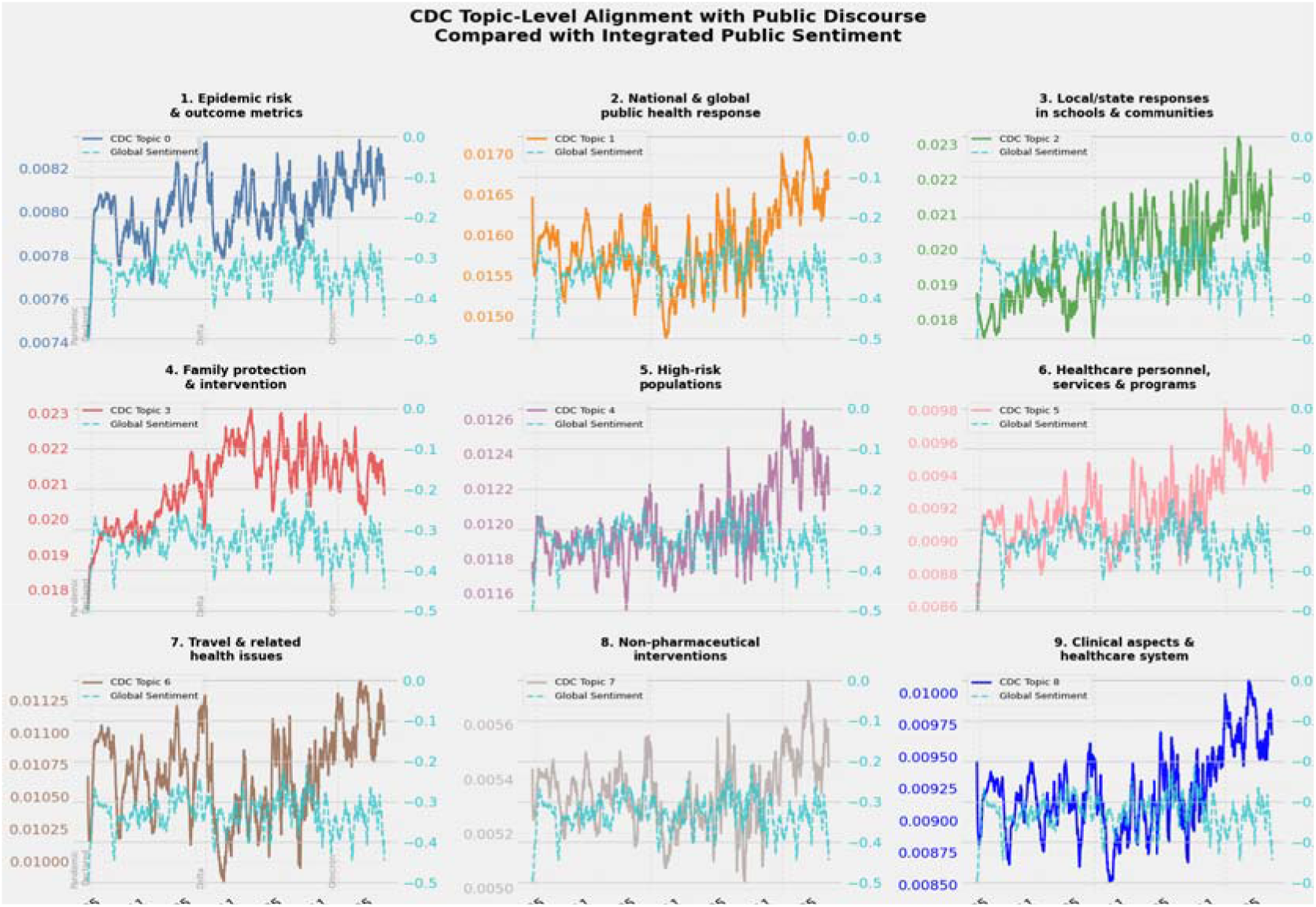
Topic-level alignment trends for all CDC topics compared with integrated expected public sentiment over time. Solid lines: alignment between that CDC topic and general public discourse. Dashed lines: integrated expected public sentiment. If the line consistently stays below 0, then the public sentiment overall is negative and unfavorable, regardless of the alignment trend of the CDC topic.

##### Net public sentiment (*N*_*d*_)

To examine the temporal effects of CDC-public topic consistency on net public sentiment, we applied the same ARIMAX framework, incorporating lagged topic consistency as the exogeneous independent variable, autoregressive (AR), or autocorrelation, of past sentiments, and moving average (MA) effects of noises in the sentiment time series. Unlike expected public sentiment, which expresses average emotional tone, net sentiment captures the overall magnitude and intensity of emotions in public discourse. It allows us to assess the influences of topic consistency on both extremities of the sentiment and strength of public emotional responses. The results are shown in Table 5 below.

**Table 5.**
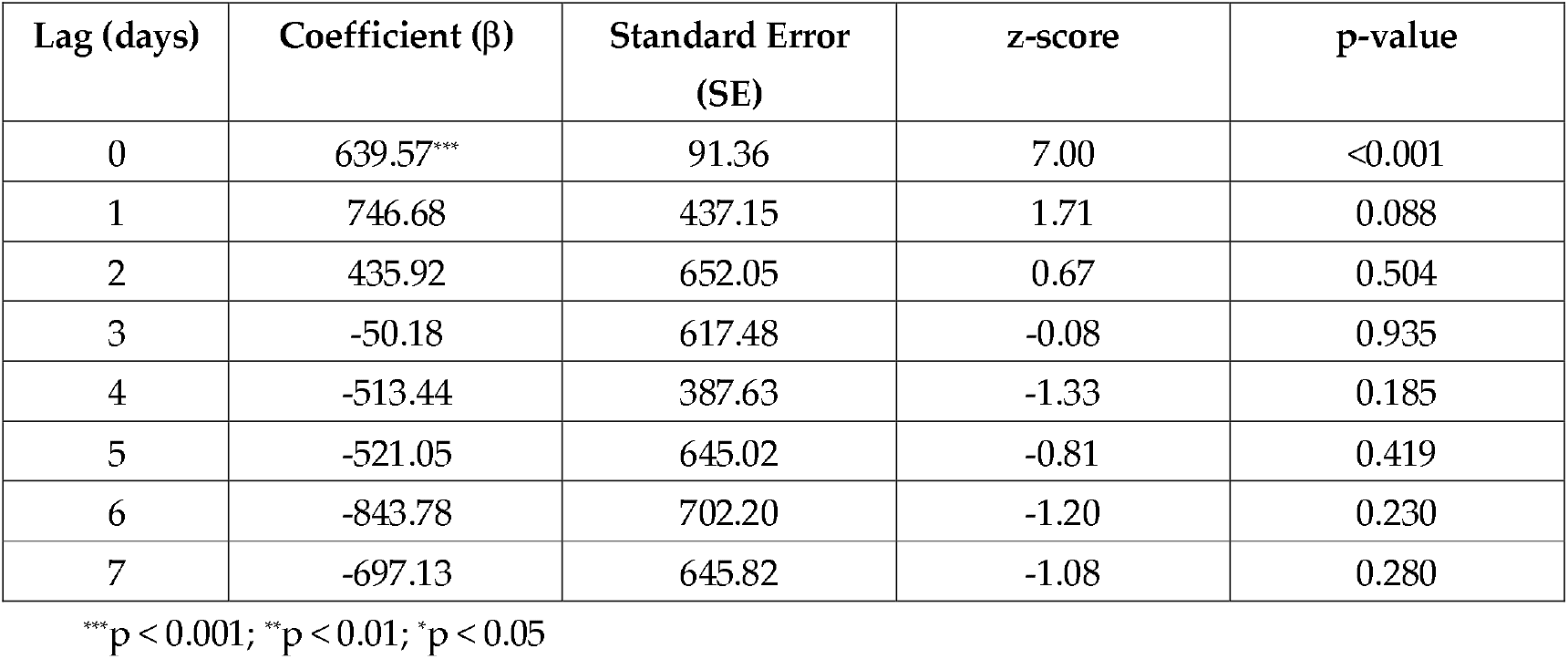
ARIMAX Results for Net Public Sentiment with Lagged Topic Consistency.

The optimal ARIMAX model order was (3, 0, 4), indicating three autoregressive (AR) terms and four moving average (MA) terms with no differencing on the time series data. ARIMAX model for the net public sentiment reveals a distinct pattern that the CDC-public topic consistency shows a strong and significant positive association with net sentiment at the same-day lag (*β* = 639.57, p < 0.001), indicating that higher topic alignment between CDC communication and general public discourse is substantially associated with increased overall emotional intensity of the public. In contrast to the expected public sentiment, where topic alignment was persistently associated with a more negative tone, the effect of topic alignment on the overall emotional intensity is primarily immediate. Thus, public emotional intensity responds fast instead of delayed towards CDC communication.

#### 3.2.3. Overall Relationship

To examine the relationship between topic consistency and public sentiments, we compared topic consistency with both expected public sentiment (Figure 8) and net public sentiment (Figure 9) over time. Figure 8 shows that expected public sentiment, representing average emotional tone, remains consistently negative throughout the study period, despite increases in topic alignment over time. In addition, sentiment variability increases over time, particularly during later pandemic phases, indicating more unstable public emotional responses.

**Figure 8.**
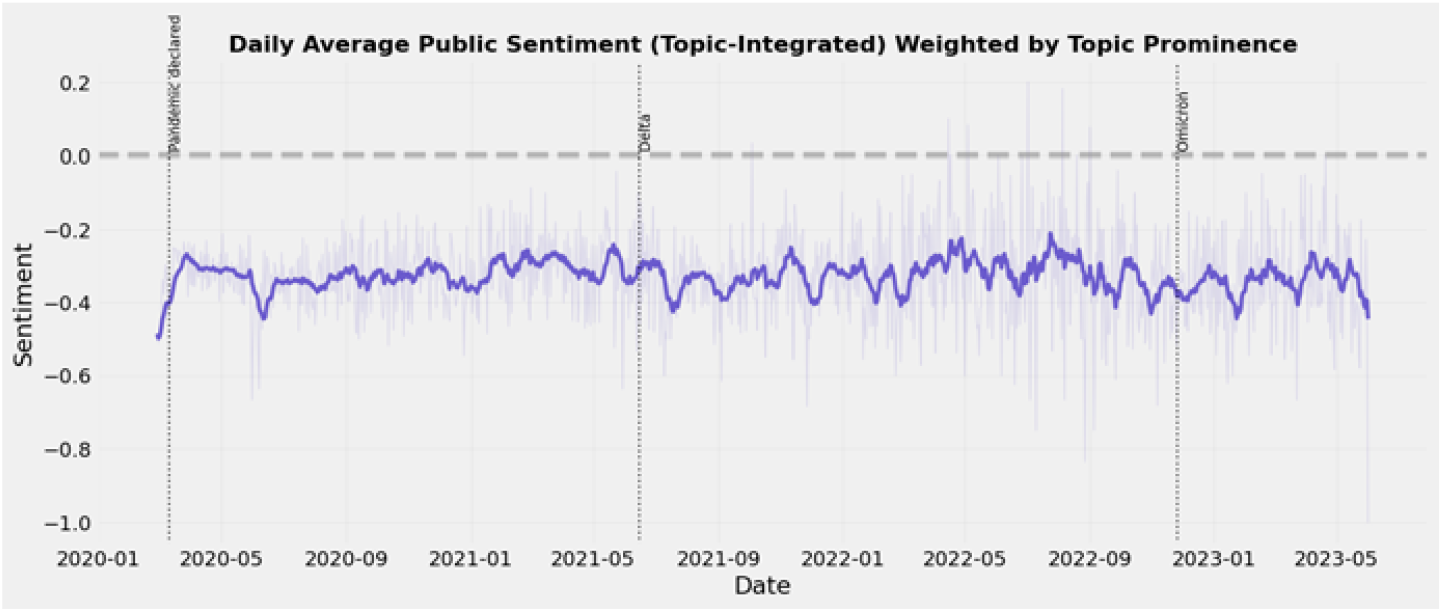
Expected public sentiment over time (topic-integrated), showing consistently negative average emotional done despite increasing topic alignment

**Figure 9.**
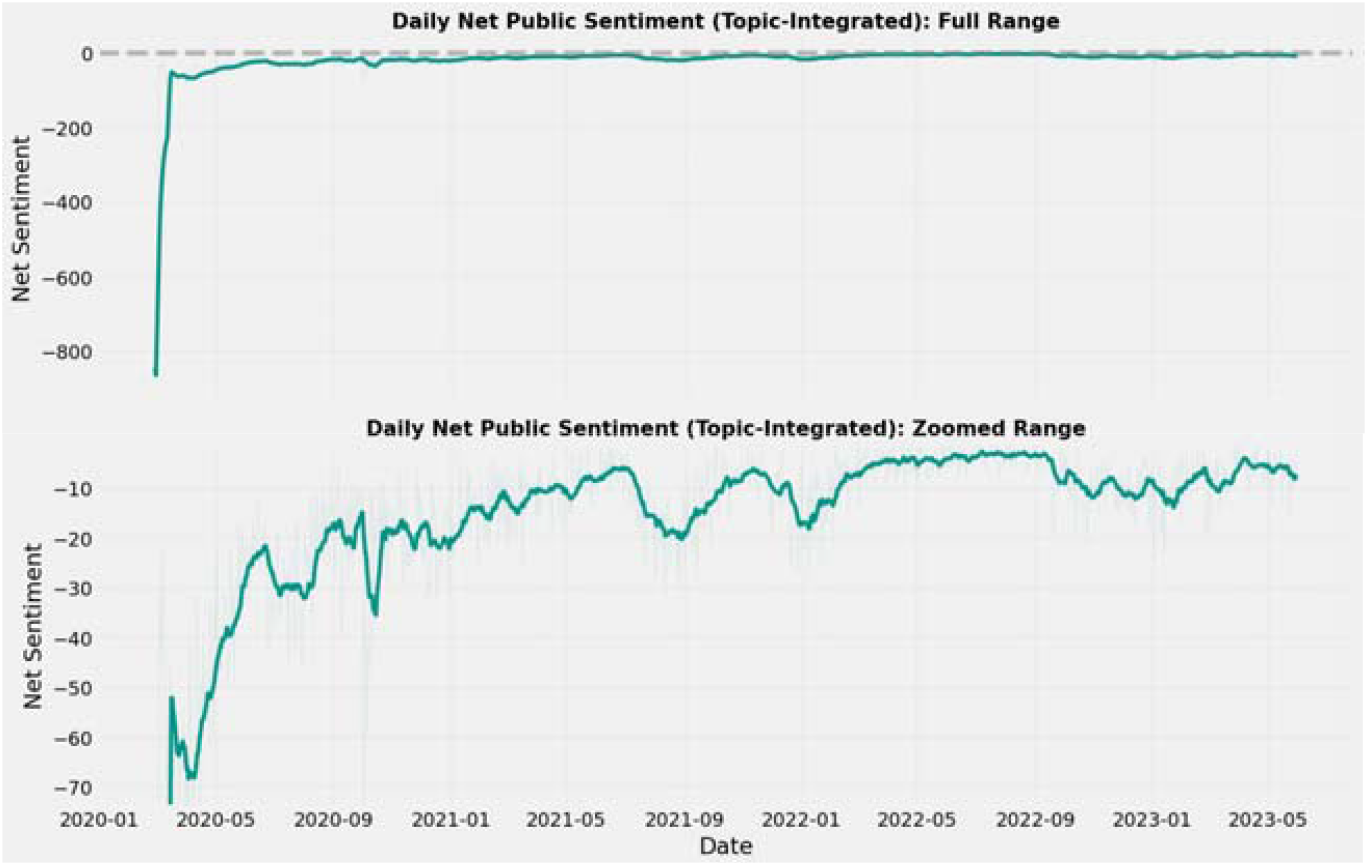
Net public sentiment over time (topic-integrated). Top panel: the full value range; bottom panel: the truncated range excluding extreme outlines (1^st^ – 99^th^ percentile) to highlight overall trends.

For net public sentiment, which captures overall emotional intensity, we visualized its time series in both full value range and a truncated range excluding extreme outliers (1^st^ – 99^th^ percentile) to better examine the overall sentiment dynamics without the extreme values. As shown in Figure 9, net public sentiment shows strong fluctuations, particularly during early pandemic phases, indicating periods of highly concentrated emotional intensity. The full-range visualization, presented on top panel, captures extreme values, while the truncated range visualization in the bottom panel exhibits temporal trends excluding the extreme outliers. Net public sentiment initially shows a sharp decline during the early pandemic phase, followed by gradual stabilization over time, indicating that the public emotional responses were negative, concentrated and intense during the early pandemic.

Early pandemic stages reflect time periods where multiple public discourse topics were simultaneously strongly negative, resulting in a substantial imbalance in sentiment polarity, were negative sentiment significantly outweighed positive sentiment. For example, on February 29, 2020, CDC and Washington Department of Public Health report the first death of an individual with laboratory-confirmed COVID-19 in the U.S. [23], net sentiment reached its lowest value, reflecting an intense negative public sentiment. During this period, the topic related to political leaders and national response (General Public Topic 4 in Table 1), characterized by keywords such as “president”, “response”, “death”, and “crisis” consistently exhibited strong negative sentiment relative to other general public discourse topics. This suggests that public discourse about leadership and national response to the crisis contributed to heightened overall emotional intensity during early pandemic phases. These results suggest that increasing topic alignment is associated with persistently negative average emotional tone and periods of intensified emotional responses.

## 4. Discussion

### Principal Findings

This study developed and applied a novel end-to-end infoveillance framework to examine the dynamic relationship, particularly topic consistency and public sentiments, between CDC communications and general public discourse on social media during the COVID-19 pandemic. Two large-scale COVID-19 Twitter/X datasets were comprehensively collected and analyzed: 17,524 CDC tweets from December 2019 to January 2022, and 67,895 general public discourse tweets from February 2020 to June 2023. Using the Biterm Topic Modeling (BTM), we identified 9 distinct CDC communication topics and 10 general public discourse topics, representing varying topic themes in public discussions and official health agencies’ communications. While they all address COVID-19-related content, they reveal distinct focus of topics in the two corpora. The general public discourse topics captured a broad spectrum of public opinions and concerns related to COVID-19, including economic impact, politics, sports and leisure, po- and state impact, media and communications, personal impact, and spread and protection. CDC health communication topics were more focused on epidemic risk and outcome metrics, national and global public health response, local and state responses in schools and communities, family protection and intervention, high-risk populations, healthcare personnel and services, travel-related health issues, non-pharmaceutical interventions, and clinical aspects and healthcare system. These results distinguished the emphases on the pandemic from both institutional and broad public lived-experience perspectives.

Second, a key contribution of this study is the development of a novel, universal topic consistency scoring system, which is able to quantify the dynamic alignment or divergence between two sets of continuous discourse samples, for instance, general public discourse and CDC communications over time in this study. The daily topic consistency scores measure how closely CDC communication topics align with general public discourse topics, weighted by their daily prominence. By integrating semantic similarity with daily topic prominence, computed as weights, this approach reflects the alignment in the context of what the public is actually discussing at a given time point. The findings show that their alignment patterns vary across CDC topics and evolve over time. Some topics (e.g., local and state responses, and family protection) showed consistently stronger alignment with public discourse, while others (e.g., healthcare services and non-pharmaceutical interventions) showed lower and more fluctuating alignment over time, indicating differences in how public interests vary and shift in specific official health communication topics.

Third, this study introduces two complementary public sentiment measures, expected sentiment and net sentiment, to comprehensively quantify public sentiment from different perspectives. Expected sentiment reflects how people feel on average, i.e., the average emotional tone, where net sentiment reflects how strongly those emotions are expressed collectively, i.e., overall emotional intensity. Our findings show that expected sentiment remained consistently negative over time, even during periods of higher topic alignment with the CDC communications. This suggests that stronger public topic alignment does not necessarily indicate agreement or positive public reception; instead, it is likely that the public is actively concerned about and focused on the same topics. Net sentiment showed more negative spikes, particularly during early pandemic phases, indicating the topic alignment during these periods was associated with stronger overall emotional intensity in public discourse. Together, the results reveal distinct but complementary public sentiment dynamics, showing that topic alignment reflects public concerns and attention, rather than agreement or effective communication from health agencies.

Finally, this study applied multivariate time series analysis, the autoregressive integrated moving average with exogenous variables (ARIMAX) model, to examine and quantify the temporal dynamics between topic consistency and public sentiments over time. For expected sentiment, topic consistency influenced average emotional tone both immediately and following days of official health communications, indicating lagged effects and a recurring weekly periodic pattern as well. For net sentiment, topic consistency primarily influenced emotional intensity on the same day, indicating that effects in overall emotional intensity occur immediately after the official communications. Therefore, these results show that topic consistency influences the average public emotional tone both immediately and over time, and while its influence on overall emotional intensity is primarily immediate. This highlights that different aspects of public sentiment respond to topic alignment in distinct ways.

### Limitations and Future Works

This study has a few limitations. First, the analysis is based solely on Twitter/X platform, which may not fully capture diverse user demographics. Expanding the analysis to multiple social media platforms (e.g., Instagram, Reddit) could provide a more comprehensive and representative view of general public discourse, obtaining richer and more nuanced insights into evolving public opinions and emotional responses, and help strengthen and validate the framework with a holistic multimedia structure and broader user population. Second, although the Biterm Topic Modeling and sentiment analysis are well-suited for large-scale short-text data, these methods may not fully capture subtle linguistic features such as sarcasm, ambiguity, or mixed sentiments, incorporating more advanced natural language processing approaches may improve the ability to capture these subtleties. Third, while the proposed topic consistency scoring system quantifies alignment between CDC communications and general public discourse, it does not establish causal relationships between the two.

Future work can enhance the generalizability and robustness of this infoveillance framework by incorporating data across multiple social media platforms and integrating more advanced large language models to better capture contextual nuances in public opinions and emotions. In addition, future work can explore causal relationships among official health communications (especially key intervention policies), general public discourse and responses, and observed outcomes (e.g., vaccination adoption rate and hospitalization). The modular design of this infoveillance framework also allows for the integration of alternative topic modeling approaches, similarity measures, and sentiment metrics for broader applications across different domains and contexts beyond health informatics.

## 5. Conclusions

This study presents a novel, end-to-end infoveillance framework to examine and investigate the dynamic relationship between CDC communications and general public discourse during the COVID-19 pandemic. By integrating topic modeling, topic consistency scoring system, and multivariate time series analysis modules, this framework quantifies how closely CDC official health communications align with general public discourse over time. Importantly, the findings demonstrate that increased topic alignment reflects public attention and concerns rather than agreement or positive reception from the public, especially regarding intervention policies. A topic-sentiment dynamic module, integrating the topic consistency scoring system with the multivariate time series model, was introduced, incorporating two new complementary public sentiment measures, expected public sentiment (average public emotional tone) and net public sentiment (overall public emotional intensity). Results show that changes in expected public sentiment occur both immediately and over time, whereas net public sentiment responds more immediately towards official communications. These results emphasize the importance of interpreting and understanding public engagement and reactions through topic alignment, expressed by the public and health agencies, but also through the emotional context in which that alignment occurs.

Overall, this framework provides a flexible and generalizable approach for understanding the dynamic associations and evolving relationship between CDC communications and general public discourse during different stages of the COVID-19 pandemic. It offers meaningful insights for more effective communication between health agencies and the public during future public health emergencies.

## 6. Patents

This section is not mandatory but may be added if there are patents resulting from the work reported in this manuscript.

## Supplementary Materials

The following supporting information can be downloaded at: https://www.mdpi.com/article/doi/s1, Figure S1: title; Table S1: title; Video S1: title.

## Author Contributions

For research articles with several authors, a short paragraph specifying their individual contributions must be provided. The following statements should be used “Conceptualization, X.X. and Y.Y.; methodology, X.X.; software, X.X.; validation, X.X., Y.Y. and Z.Z.; formal analysis, X.X.; investigation, X.X.; resources, X.X.; data curation, X.X.; writing—original draft preparation, X.X.; writing—review and editing, X.X.; visualization, X.X.; supervision, X.X.; project administration, X.X.; funding acquisition, Y.Y. All authors have read and agreed to the published version of the manuscript.” Please turn to the CRediT taxonomy for the term explanation. Authorship must be limited to those who have contributed substantially to the work reported.

## Institutional Review Board Statement

Not applicable

## Informed Consent Statement

Not applicable

## Data Availability Statement

The original CDC data in the study are openly available on Twitter/X. The original general public discourse data in the study were obtained from 3^rd^ party platform, Brandwatch and are available https://www.brandwatch.com/ with the permission of Brandwatch.

## Acknowledgments

This study is supported by the NSF grant DMS-2436227.

## Abbreviations

The following abbreviations are used in this manuscript:

MDPI: Multidisciplinary Digital Publishing Institute
DOAJ: Directory of open access journals
TLA: Three letter acronym
LD: Linear dichroism

## Disclaimer/Publisher’s Note

The statements, opinions and data contained in all publications are solely those of the individual author(s) and contributor(s) and not of MDPI and/or the editor(s). MDPI and/or the editor(s) disclaim responsibility for any injury to people or property resulting from any ideas, methods, instructions or products referred to in the content.

