## Supplementary Materials for "Dynamic Topic Alignment and Sentiment between Official Health Communication and General Public Discourse during COVID-19: A Comprehensive Infoveillance Framework"

**Data Collection**

**CDC Communication Tweets**:

**Search query used to obtain CDC tweets for data collection**:

(ncov OR ncov-19 OR sars OR SARS-COV-2 OR "corona virus" OR pandemic OR pheic OR "wuhan virus" OR "china virus" OR "wuhan pneumonia" OR "wuhan flue" OR kungflue OR covid19 OR covid OR "covid 19" OR coronavirus OR vaccine OR vaccines OR vaccination)

**Data collection process of CDC tweets**:

Using the Twitter academic API and search query, we retrieved a total of 17,524 English tweets posted by seven official CDC-affiliated Twitter accounts from December 7, 2019, to January 15, 2022. These accounts are: @CDCgov: CDC’s official Twitter source for daily credible health and safety updates from Centers for Disease Control & Prevention; @CDCEmergency: CDC Emergency, which tweets ways to for public health preparedness during emergency responses; @CDCDirector: account of the CDC director of the time; @CDCGlobal: CDC Global health, which tweets about how CDC strives to contribute to saving lives, reducing disease, and improving global health around the world; @CDCtravel: CDC Travel health, which voices to help travelers and their clinicians prevent illness and injury during international travel; @DrKhabbazCDC: past Twitter account from former director of CDC emerging infections (NCEZID), Dr. Rima Khabbaz. NCEZID works to protect people from emerging and zoonotic infectious diseases, from anthrax to Zika; @CDCMMWR: MMWR (Morbidity and Mortality Weekly Reports), which is CDC’s primary vehicle for scientific publication of timely, authoritative, and useful public health information and recommendations. We consolidated these tweets into a data frame of daily tweet counts of each of the seven CDC-affiliated accounts during the time period. The associated metadata include tweet posting dates, textual data of the tweets, tweet account ID, account types (organization or individual), public engagement metrics (e.g., number of retweets, replies, likes, and quotes), referenced tweet type (retweeted, replied to, and quoted), and referenced tweet IDs.

**General Public Discourse**:

This study is based on a dataset of highly engaged tweets related to general public discourse on COVID-19, collected using the BrandWatch platform. BrandWatch provides access to social media content through customizable, keyword-driven queries. We focused on tweets with an engagement score of 10 or higher, ensuring that the dataset represents content that elicited notable public attention.

The engagement score on Twitter (now X) is calculated as the sum of a tweet’s likes, reposts (retweets), and replies. In BrandWatch, this is a network-specific "sum-all" metric, each platform aggregates interactions across its own set of engagement indicators. For Twitter, the formula is as follows:

*Engagement Score (Twitter) = Likes + Retweets + Replies*

This measure reflects a tweet’s overall interaction level and filters out low-visibility or low-impact posts.

To gather relevant content, we used a broad keyword-based query encompassing medical, colloquial, and politicized references to COVID-19 (Table 1). The keywords were searched in either the bodies or the  titles of the tweets, and results were filtered to include only those from Twitter (site:twitter.com).

**Table 1.** COVID-19 Keywords Used in Query

| Category | Keywords |
| --- | --- |
| General Keywords | ncov, ncov-19, sars, SARS-CoV-2, coronavirus, pandemic, pheic |
| Politicized Terms | "wuhan virus", "china virus", "wuhan pneumonia", "wuhan flu", kungflue |
| Standard COVID Terms | covid19, "covid-19", covid, "covid 19" |
| Official Terminology | "Public Health Emergency of International Concern" |

The data collection period spans the full timeline of the COVID-19 pandemic, with the query covering from December 7, 2019, to May 31, 2023. However, the actual range of highly engaged tweets begins on February 28, 2020, at 00:00:06 (UTC) and ends on June 1, 2023, at 01:40:01 (UTC). A total of 67,895 tweets meeting the engagement criteria were retrieved within this timeframe. To provide temporal context, Figure 1. summarizes the number of highly engaged tweets by month.

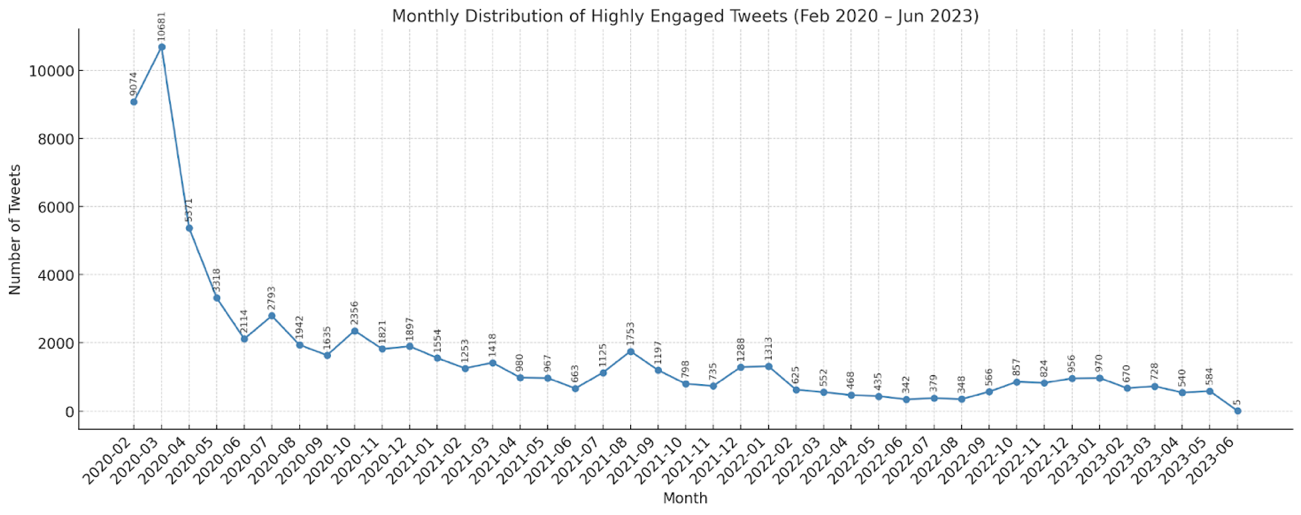

Figure 1. Monthly Distribution of Highly Engaged Tweets (UTC)

To further characterize the dataset, we computed basic descriptive statistics on the length of tweet texts, measured in words (Table 2).

**Table 2.** Descriptive Statistics of Tweet Word Counts

| **Statistics** | **Value (words)** |
| --- | --- |
| Total number | 67,985 |
| Mean | 30.72 |
| Max. | 105 |
| Min. | 1 |
| Q1 (25^th^ percentile) | 19 |
| Q2 (Median) | 32 |
| Q3 (75^th^ percentile) | 42 |
| Standard deviation | 12.96 |

This dataset provides a rich basis for analyzing public sentiment, topic trends, and engagement patterns over the course of the pandemic, with an emphasis on content that resonated widely on social media.

**Visualizations of general public discourse topics (10) plots using *tmplot*:**

Topic 0:

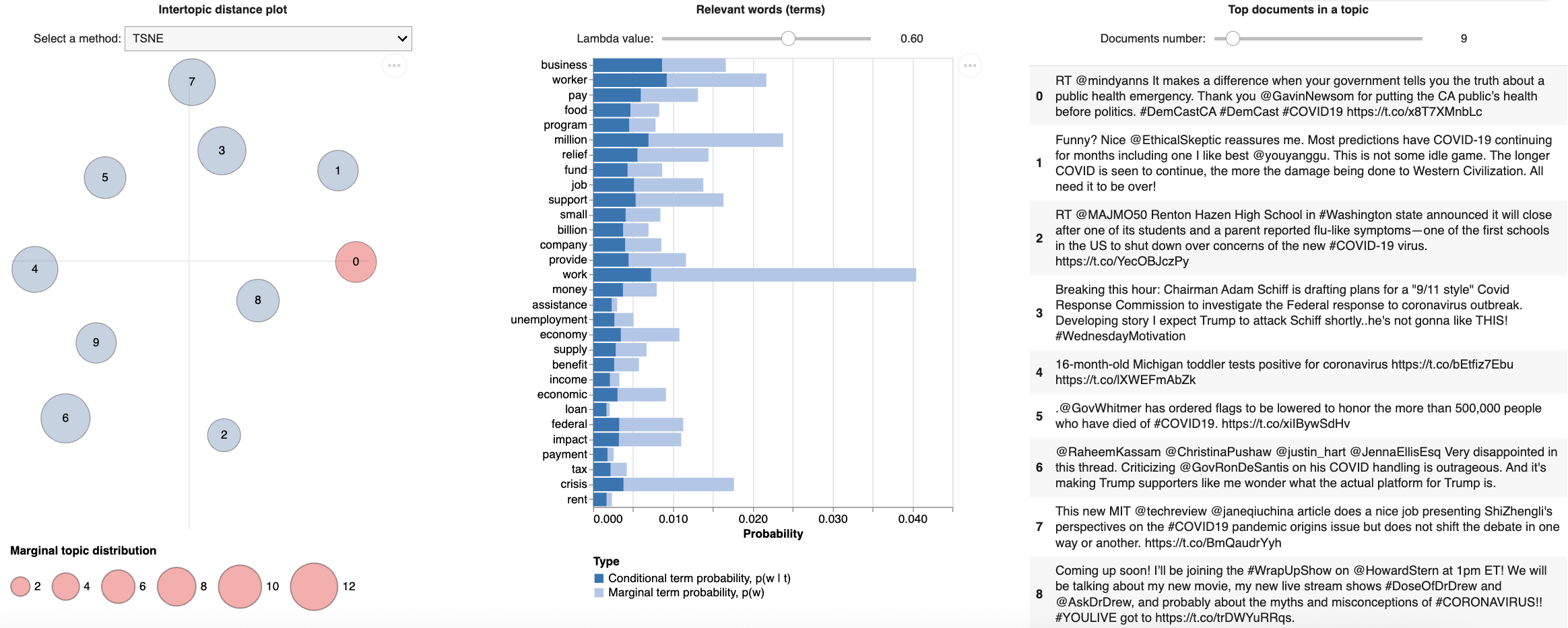

Figure 2. General Public Discourse Topic 1: Economic impact

Topic 1:

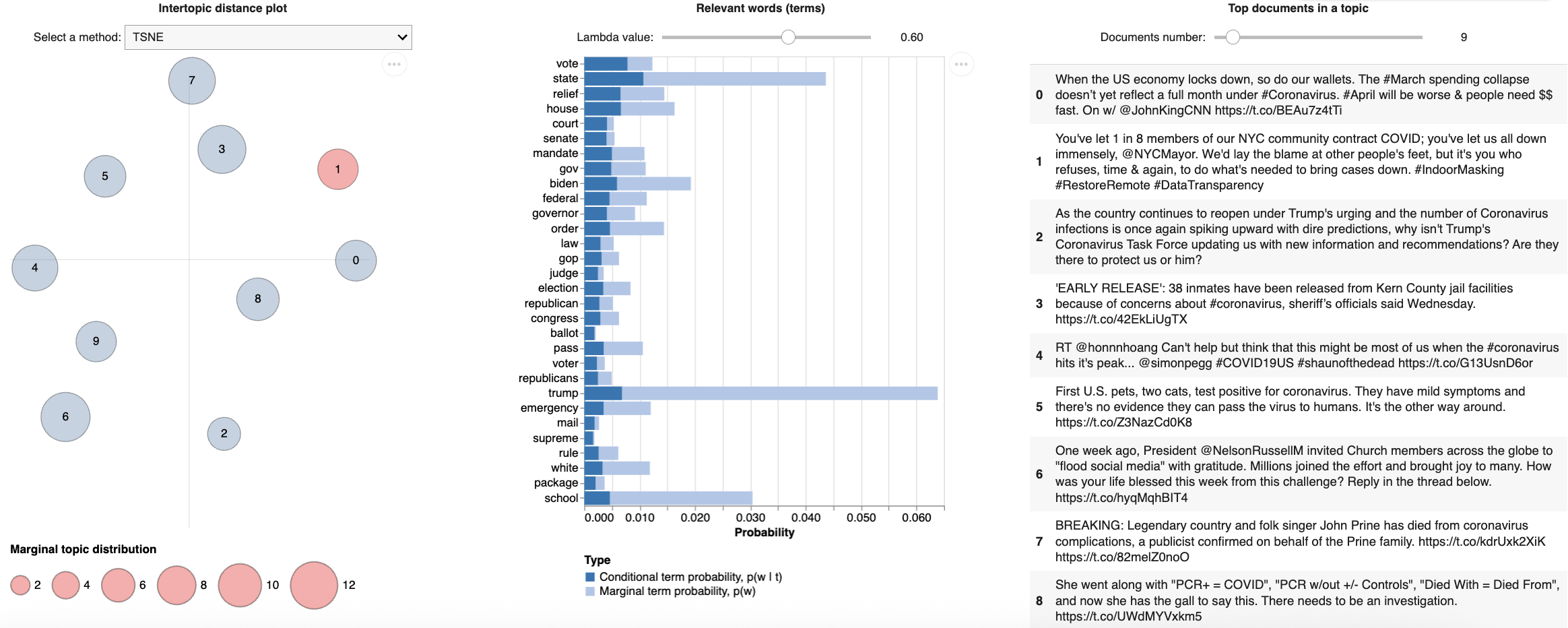

Figure 3. General Public Discourse Topic 2: Politics

Topic 2:

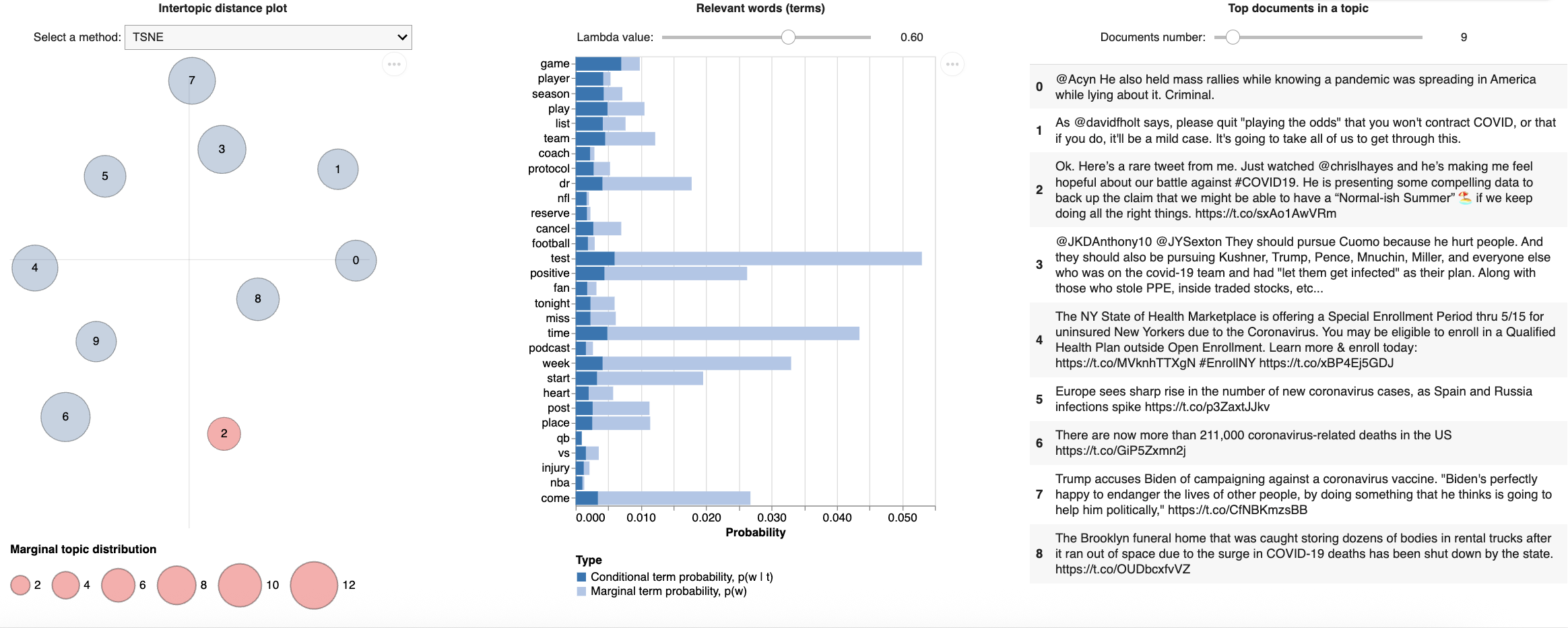

Figure 4. General Public Discourse Topic 3: Sports and leisure

Topic 3:

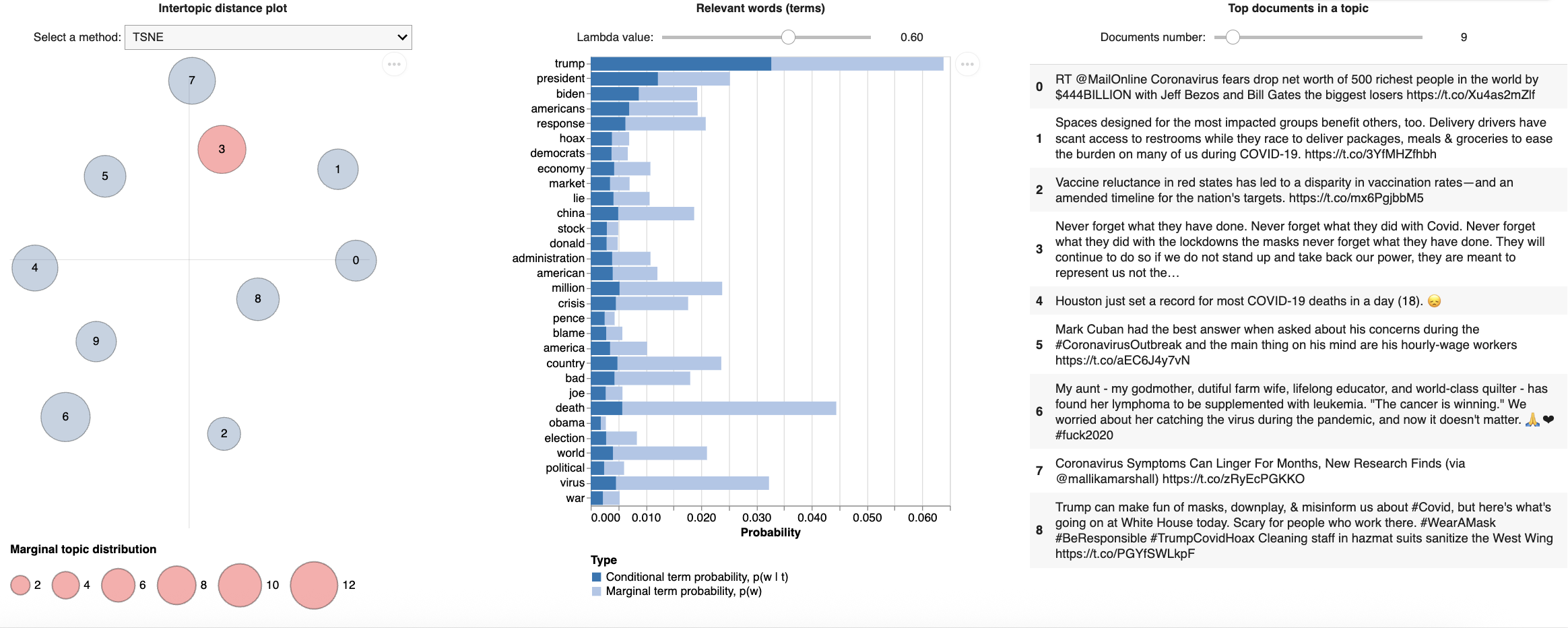

Figure 5. General Public Discourse Topic 4: Political leaders

Topic 4:

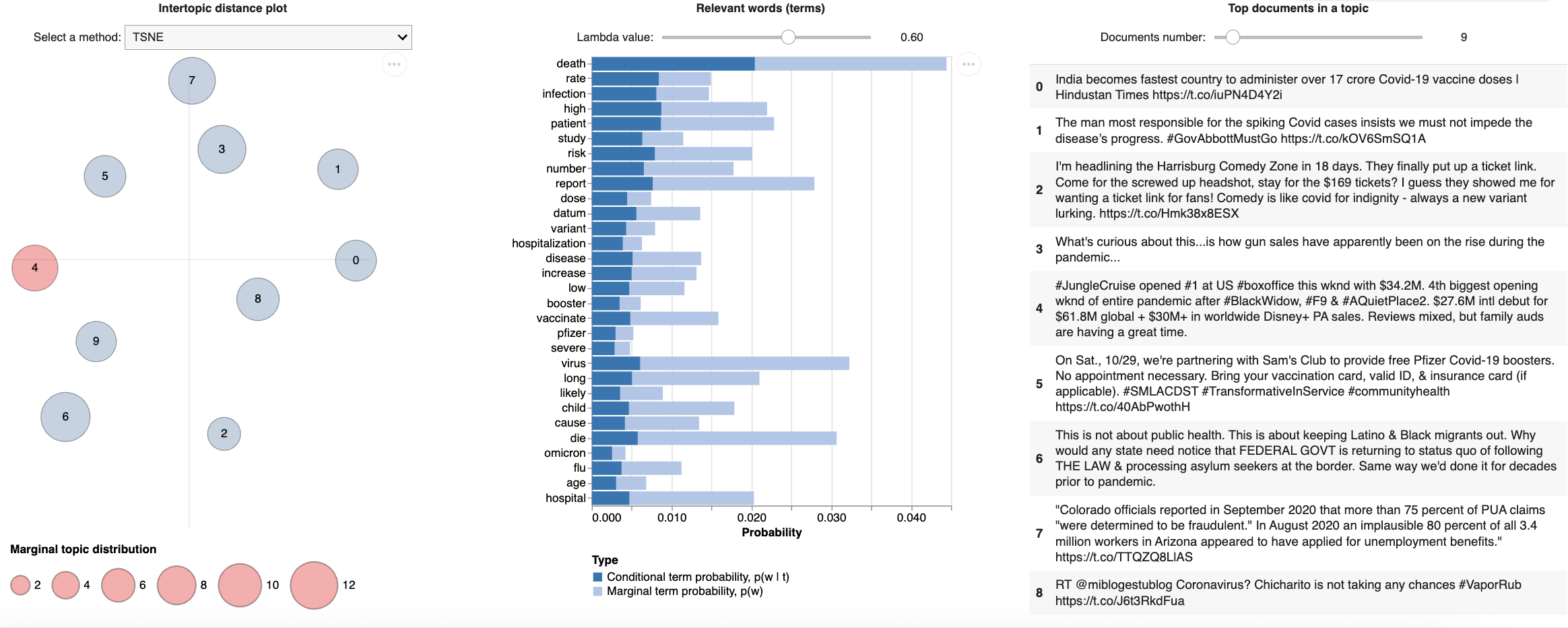

Figure 6. General Public Discourse Topic 5: Health outcomes

Topic 5:

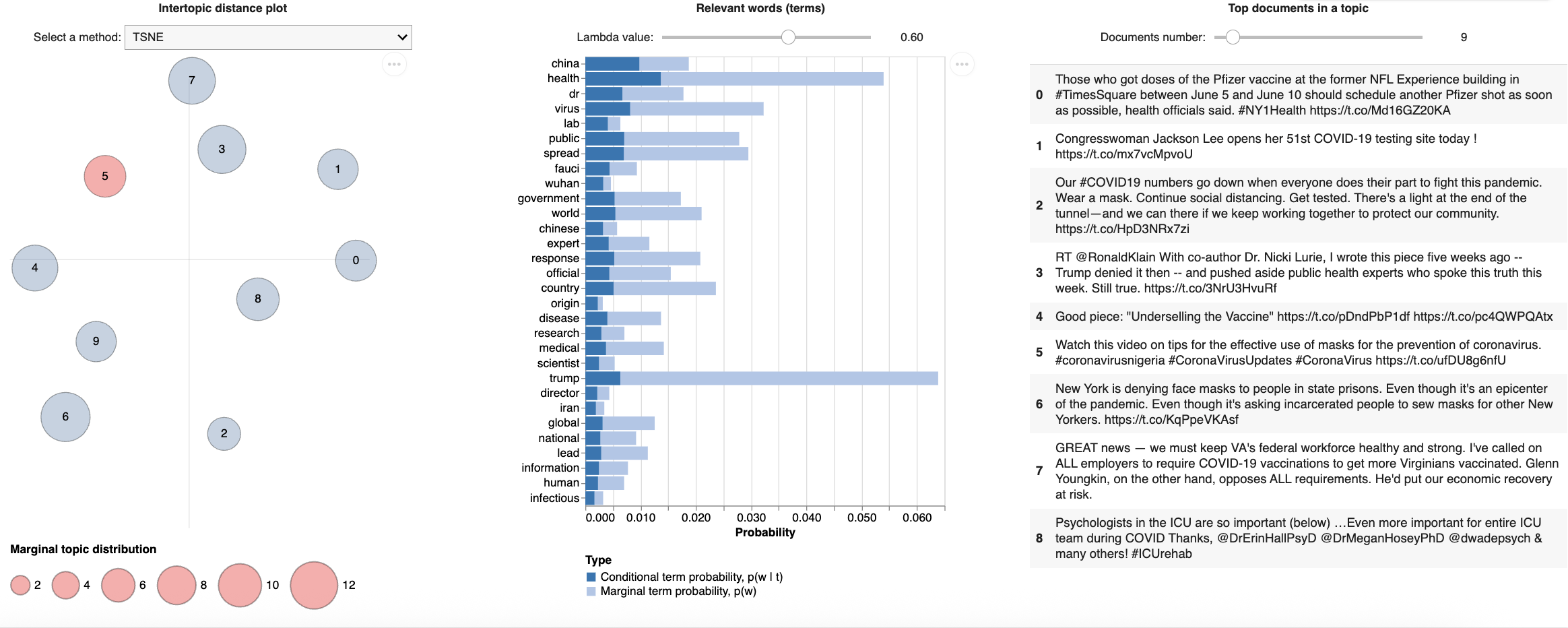

Figure 7. General Public Discourse Topic 6: Global health and relations

Topic 6:

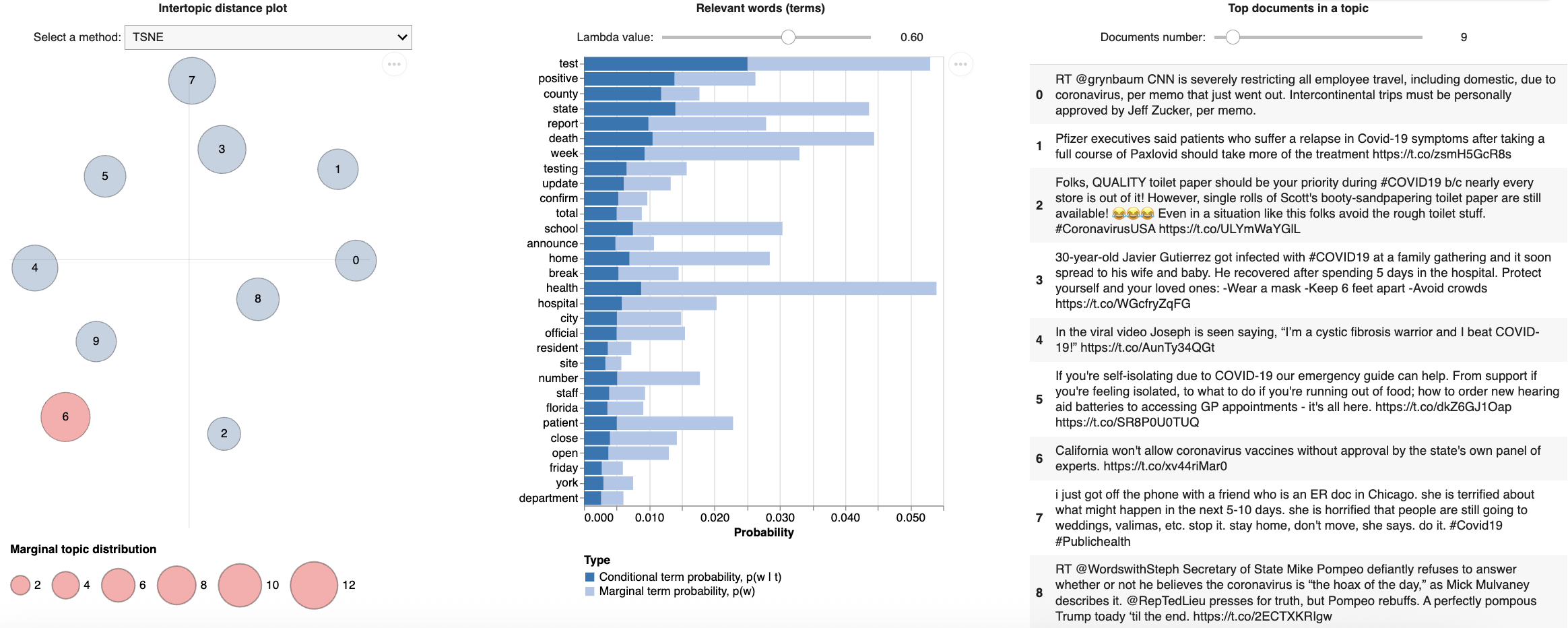

Figure 8. General Public Discourse Topic 7: Local and state impact

Topic 7:

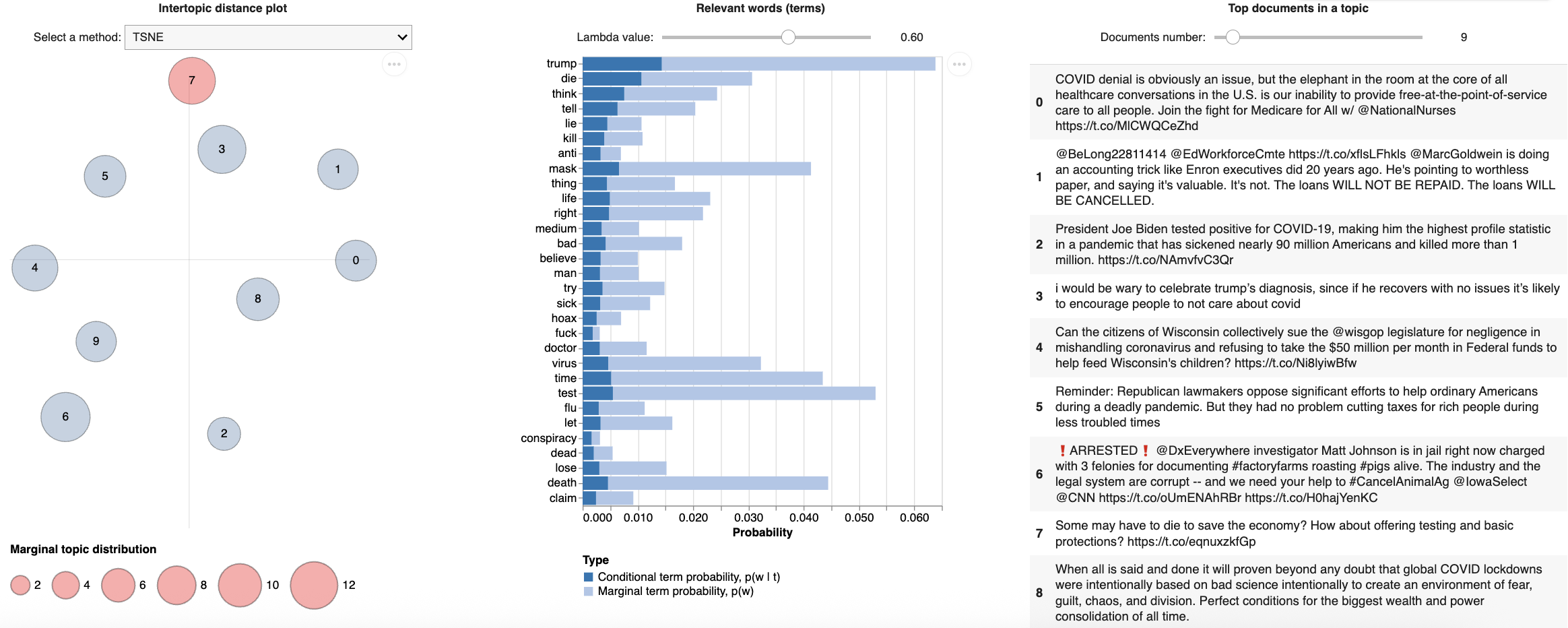

Figure 9. General Public Discourse Topic 8: Media and communications

Topic 8:

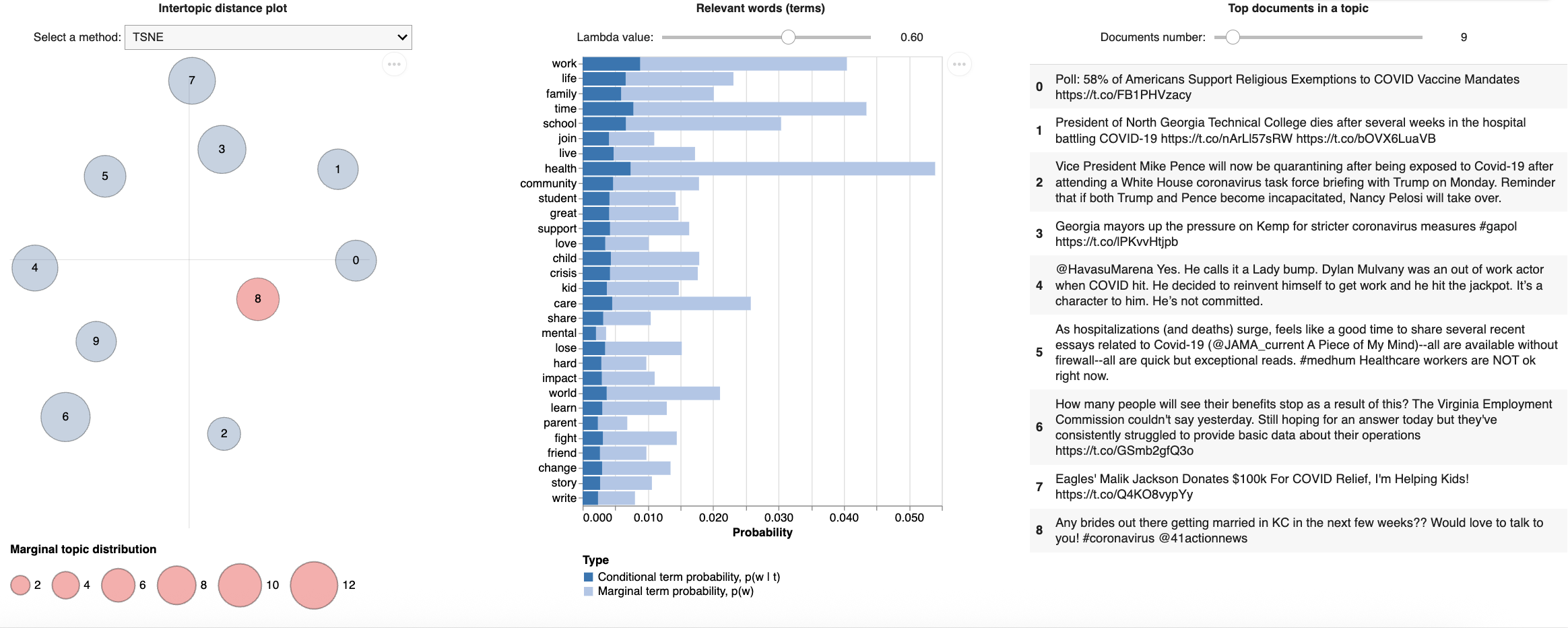

Figure 10. General Public Discourse Topic 9: Personal impact

Topic 9:

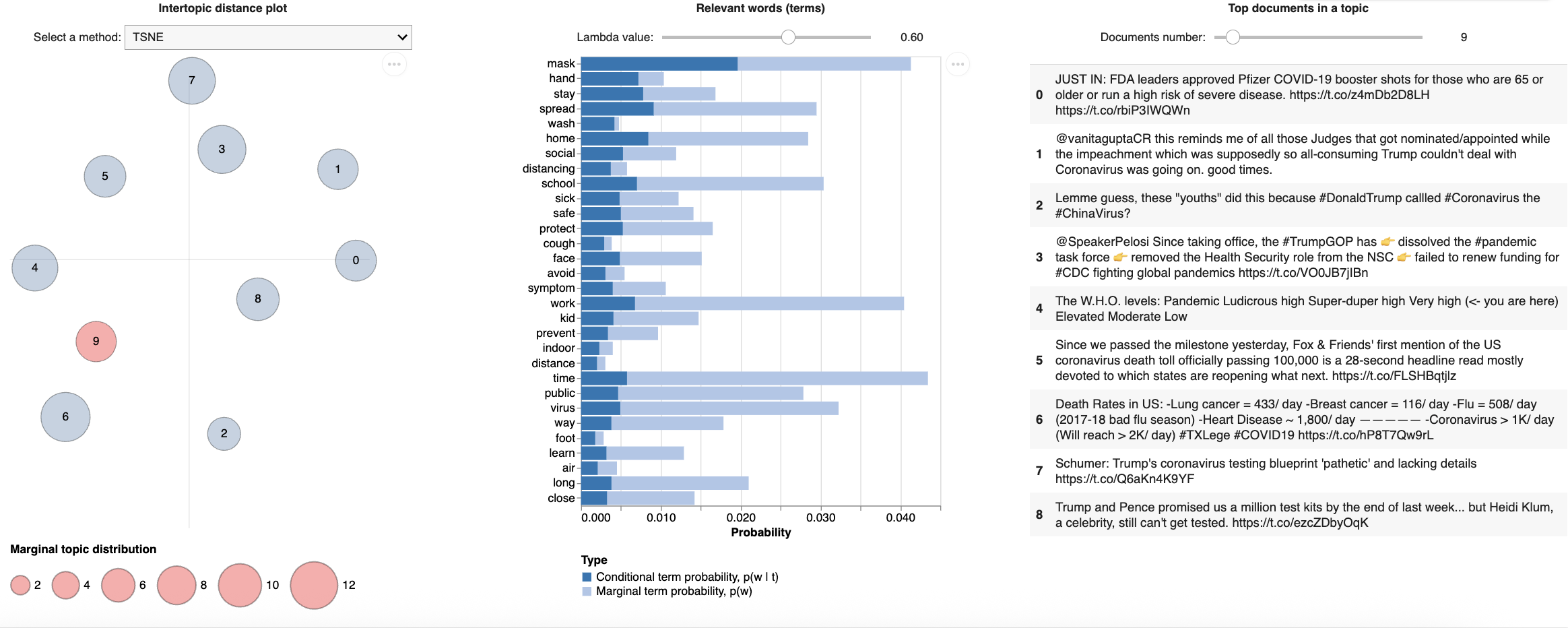

Figure 11. General Public Discourse Topic 10: Spread and protection

**Autoregressive (AR) model**:

AR is a regression model where linear combinations of the past terms are the predictors of the variable in the future. Assuming the values of the variable are independently and normally distributed, AR(*p*) model of order or lag *p* can be written as:

y_t_ = *c* + *φ*_1_y*_t_*_-1_ + *φ*_2_y*_t_*_-2_ + … + *φ_p_*y*_t_*_-_*_p_* + ε*_t_* = *c* + $\sum_{i=1}^{p} \varphi_{i}y_{t-i}$ + ε*_t_* [24],

or in a simplified version with backward shift operator form:

*φ*_p_(B)y_t_ = 1 - *φ*_1_B + … + *φ_p_*B_p_ [25],

where *c* = average of changes between the sequential observations,

ε*_t_* = white noise,

y*_t_* = lagged values, or predictor values of the next value of the variable y.

**Moving Average (MA) model**:

MA is another linear model of past forecast errors in a univariate time series. MA(*q*) model of order or lag *q* can be written as:

y*_t_* = *c* + *θ*_1_ε*_t_*_-1_ + *θ*_2_ε*_t_*_-2_ + … + *θ_q_*ε*_t_*_-_*_q_* + ε*_t_* = *c* + $\sum_{j=1}^{q} \theta_{j}y_{t-j}$ + ε*_t_* [26],

or in backward shift operator form:

*θ_q_*(B)y*_t_* = 1 - *θ_1_*B + … + *θ_q_*B*_q_* [25].

**Autoregressive integrated moving average (ARIMA) model**:

ARIMA model consists of both AR(*p*) and MA(*q*) as well as order *d* differencing term, resulting in the following ARIMA (*p*, *d*, *q*) model:

y′_t_ = *c* + *φ*_1_y′*_t_*_-1_ + … + *φ_p_*y′*_t_*_-_*_p_* + *θ*_1_ε*_t_*_-1_ + *θ*_2_ε*_t_*_-2_ + … + *θ_q_*ε*_t_*_-_*_q_* + ε*_t_* [27],

or in backward shift form:

(1 - *φ*_1_B + … + *φ_p_*B_p_)∇^d^y*_t_* = *c* + (1 + *θ*_1_B + … + *θ*_q_B^q^)ε*_t_* or *φ*(B) )∇^d^y_t_ = *c* + *θ*(B)ε*_t_* [28] (see supplementary materials for details on the parameters),

where y′_t_ = time series after differencing, which is presented as (1 – B)^d^y*_t_* as d differences in backward shift operator form,

∇^d^ = (1 – B)^d^y*_t_*,

*p* = order of past lagged values of for each time point of autoregressive model,

*d* = the degree of differencing occurred, or the number of times performs integration,

*q* = order past lagged errors for the error term as a combination of predictors of moving average model.

**Biterm Topic Model (BTM)**

Biterm topic model (BTM) is a generative probabilistic model designed to extract latent topics from short and sparse texts by directly modeling the biterms as word co-occurrences at the corpus level. A biterm is an unordered pair of words that co-occur in a short text, and BTM extracts these biterms across the whole corpus to learn the global topic distributions.

The specific generative process of the corpus in BTM with diagram is shown below [1]:

Notations:

K: number of topics in the corpus

*z*: A topic assignment from the corpus;

*θ*: Multinomial global topic distribution of the whole corpus;

*φ_z_*: Multinomial topic-word distribution for topic *z*;

*α*, *β*: Dirichlet priors, or hyperparameters, for *θ* and *φ_z_*, respectively;

*b* = (*w_i_*, *w_j_*): A biterm drawn with an unordered word pair from the corpus;

B: The set of all biterms

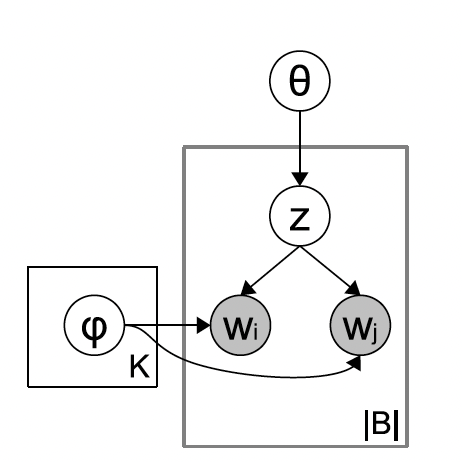

1. For each topic z ∈ {1, …, K};

- Draw a topic-specific word distribution *φ*, where *φ_z_* ~ Dir(*β*)

2. For the entire corpus:

- Draw a global topic distribution *θ*, where *θ* ~ Dir(*α*)

3. For each biterm b = (*w_i_*, *w_j_*) ∈ B:

- Draw a topic assignment *z*, where z ~ Mult(*θ*)
- Draw two words, *w_i_* and *w_j_*, independently from the topic, where *w_i_*, *w_j_* ~ Mult(*φ_z_*)

**Rényi entropy**

By computing the Rényi entropy with different number of topics and locating the minimum entropy value for topic modeling, we are able to select the optimal number of topics that generates the maximum information with distinct topics and is the closest to the real latent topic structure.

Rényi entropy is a generalization of Shannon entropy and is used to measure the “density of states” of a topic-word distribution in topic modeling [2] while varying the number of topics and hyperparameters. Lower entropy indicates a more concentrated or cohesive topic that is close to the latent structure, and higher entropy indicates a more diffused topic.

In topic modeling, the Rényi entropy is defined as follows [3]:

$S_{Q}^{R}$ = $\frac{ln \left( Z_{q} \right)}{q-1}$ = $\frac{qln\left( q\tilde{P} \right)+ q^{-1}ln\left( \tilde{\rho} \right)}{q-1}$,

where $q$= $\frac{1}{T}$, *T* is the number of topics, $\tilde{\rho}$ = $\frac{N}{WT}$ is the “density-of-states” function that represents the high-probable word-topic proportions in the corpus [4], that higher $\rho$ implies more focused or cohesive topics, W is the number of vocabulary or unique words in the data, N is the number of words with high probability, $\tilde{P}= \frac{1}{T}\sum_{wt} \phi_{wt}\boldsymbol{1}_{\{\phi_{wt}>\frac{1}{W}\}}$ is the sum of probabilities of all words of high probability, an indicator function where value of the function is 1 if $\phi_{wt}\geq$ $\frac{1}{W}$, 0 if $\phi_{wt}<$ $\frac{1}{W}$ . The optimal number of topics and the optimal values of hyperparameters indicate the minimum Rényi entropy, i.e., as the number of topics becomes optimal, the word-topic probability distribution is the most informative, resulting in the minimal entropy value and high semantic resolution [2].

**Cosine Similarity:**

Cosine similarity measures the how similar two vectors of an inner product space, or the dot product for Euclidean space [6]. It can be obtained by computing the cosine of the angle between the two vectors to determine if the two vectors are pointing in the same direction. It can be used to measure the similarity between documents in text analysis. The cosine similarity between two vectors is defined as:

Cosine similarity (*x*, *y*) = $sim(x,y)$ = $\frac{x \cdot y}{\left| \left| x \right| \right| \left| \left| y \right| \right|}$ [6],

where $x\cdot y$ is the dot product of the *x* and *y* vectors: $x\cdot y$ = $\sum_{i=1}^{n} x_{i}y_{i}$;

$\left| \left| x \right| \right|$ is the Euclidean norm of vector ***x*** = ($x_{1},x_{2}$,…, $x_{p}$), and is defined as $\sqrt{x_{1}^{2}+ x_{2}^{2}a+\ldots+x_{p}^{2}}$, where *p* is the total number of objects in the vector;

$\left| \left| y \right| \right|$ is the Euclidean norm of vector ***y*** = ($y_{1},y_{2}$,…, $y_{p}$), and is defined as $\sqrt{y_{1}^{2}+ y_{2}^{2}a+\ldots+y_{p}^{2}}$;

When cosine similarity value is 0, it means that vectors ***x*** and ***y*** are orthogonal of each other and they do not share similarity; the closer cosine similarity value is to 1, the smaller the angles of between the two vectors, thus the greater similarity between them.
